# Effects of *Shigella* diarrhea with and without antibiotic treatment on linear growth: an individual patient data meta-analysis of five multisite studies among children in low-resource settings

**DOI:** 10.64898/2026.07.10.26357688

**Authors:** Elizabeth T. Rogawski McQuade, Allison Codi, Patricia B. Pavlinac, Erika Feutz, Karen Kotloff, James A. Platts-Mills, David Benkeser

## Abstract

**Background:** Quantifying the effect of *Shigella* diarrhea with and without antibiotic treatment on linear growth faltering is critical to understanding the potential impact of *Shigella* vaccines.

**Methods:** Using individual-level data from five multisite studies, we estimated the effect of *Shigella* diarrhea on length/height-for-age z-score (HAZ) 60-90 days after the episode compared to diarrhea episodes with no etiology identified and non-diarrheal controls. Effects were estimated under treatment with and without antibiotics using augmented inverse probability weighted estimators with ensemble machine learning.

**Findings:** Among 26,752 diarrhea episodes, 5,503 (20.6%) were attributed to *Shigella* and of those, 2,567 (46.6%) were treated with guideline recommended antibiotics. Compared to other diarrhea episodes, *Shigella* diarrhea without treatment with guideline recommended antibiotics was associated with small reductions in HAZ (HAZ difference: -0.03, 95% CI: -0.05, -0.01), but not when treated with guideline recommended antibiotics (HAZ difference: -0.01, 95% CI: -0.02, 0.01). Compared to non-diarrheal controls, *Shigella* diarrhea was associated with decrements in HAZ in the following 60-90 days regardless of antibiotic treatment (HAZ difference overall: -0.08, 95% CI: -0.10, -0.07). A larger impact of *Shigella* diarrhea was observed among younger children.

**Interpretation:** *Shigella* diarrhea was associated with short-term decrements in height. Treatment with guideline recommended antibiotics prevented the impact of *Shigella* on linear growth compared to other diarrhea episodes but not compared to non-diarrheal controls. These results suggest that improved recognition and treatment or prevention of *Shigella* could improve child growth.

**Funding:** This work was supported by the Gates Foundation (INV-071320 to ERM and DB) and NIH/NIAID (R01AI185140 to ERM).

**Research in context:** *Evidence before this study:* We searched PubMed on June 1, 2026 for articles published in any language since Jan 1, 1990, using the search terms (shigell*)) AND ((pediatric OR paediatric OR infant* OR child*))) AND ((grow* OR linear OR height OR length OR stunt*)). We identified 436 publications, of which 8 research articles and two systematic reviews investigated the association between *Shigella* diarrhea and child growth. The studies were of different designs and employed a wide range of statistical analyses. Only one study considered the impact of antibiotic treatment on the association between *Shigella* and linear growth, and none appropriately accounted for antibiotic treatment as a mediator of the effect of *Shigella* on growth. No pooled estimates derived from a meta-analysis of the evidence were available.

*Added value of this study:* This study leverages harmonized diagnostic and covariate data from more than 26,000 episodes of diarrhea to conduct an individual patient data meta-analysis of the effect of *Shigella* diarrhea on short-term linear growth. The analysis appropriately accounts for antibiotic treatment as a mediator of the effect and considers comparisons to other diarrhea episodes and to non-diarrheal controls. Flexible modeling methods using machine learning and doubly robust estimators minimize bias and maximize the precision of estimates. *Shigella*-attributed diarrhea was associated with small but significant decrements in HAZ in the short-term, with larger effects observed among younger children. Treatment with guideline recommended antibiotics reduced the impact of *Shigella* compared to other diarrhea episodes, but did not prevent effects on HAZ compared to non-diarrheal controls.

*Implications of all the available evidence:* The impact of *Shigella* on growth faltering increases the value proposition for *Shigella-*targeted interventions. Appropriate antibiotic treatment may mitigate but will not fully prevent the effects of *Shigella* on child growth, suggesting detection and treatment of shigellosis is not sufficient to prevent growth faltering. Vaccination and other interventions for disease prevention are needed and could improve linear growth even in settings with frequent appropriate antibiotic treatment.

## Introduction

In low-resource settings, *Shigella* is a leading cause of diarrhea among children younger than 24 months,^1,2^ a critical period of child development. Nearly one-third of children living in low-resource settings experience at least one episode of *Shigella* diarrhea by age two,^3^ with an estimated incidence of 41 episodes per 100 child-years (95% CI: 38, 47).^2^ In addition to causing diarrhea and mortality, *Shigella* is associated with intestinal inflammation^3^ and is implicated in environmental enteropathy and longer-term outcomes like linear growth stunting.

Understanding the magnitude of impact of *Shigella* on linear growth faltering is critical to defining the value of *Shigella* vaccines, of which several are in or soon to be in Phase 3 trials.^4^ Concern that the relatively modest mortality burden attributed to *Shigella* will lead to limited vaccine uptake has encouraged efforts to expand the vaccine value proposition by quantifying the potential effects of *Shigella* vaccines on a broader array of outcomes, including child growth. The WHO Preferred Product Characteristics for a *Shigella* vaccine highlight that vaccine effects on stunting will be important to inform policy decisions and recommend that secondary trial outcomes should include growth faltering.^5^ Estimates from the Global Burden of Disease study for the number of diarrhea-associated DALYs among children under 5 years increased by 40% when accounting for effects on growth impairment.^6^ However, these estimates were not *Shigella*-specific. Similarly, existing modeling studies of the impact of *Shigella* vaccines on stunting assumed the effect of shigellosis on growth was equivalent to the effect of all-cause diarrhea on growth.^7,8^ Quantifying the effect of *Shigella* specifically on growth faltering would better inform projections of vaccine impact, promoting continued vaccine development and ultimately uptake by low and middle-income countries.

Importantly, these projections should consider the potential for antibiotic treatment to modify the growth impacts of *Shigella.* If antibiotics prevent *Shigella*-associated growth faltering, a vaccine may be expected to have more limited impact on growth stunting in settings where *Shigella* episodes are frequently treated with effective antibiotics. Furthermore, such evidence may support expansions to the current syndromic treatment guidelines,^9,10^ which do not recommend treatment of most cases of shigellosis that present as watery diarrhea.^2^

While prior studies have provided evidence that *Shigella* diarrhea is associated with linear growth faltering,^11–13^ differences in study design and analysis, including inconsistent incorporation of antibiotic treatment, make it challenging to compare results across studies. Using individual-level data from five multisite studies of diarrhea, we estimated the controlled direct effects of *Shigella* in the presence and absence of effective antibiotic treatment on linear growth faltering 60-90 days following the diarrhea episode. We compared *Shigella* diarrhea to episodes of diarrhea without an infectious etiology and separately compared children with *Shigella* diarrhea to non-diarrheal controls in the studies that included children without diarrhea.

## Methods

We conducted a pooled individual patient data (IPD) meta-analysis using individual-level data and harmonized covariates from five multisite studies of children with diarrhea: the Global Enteric Multicenter Study (GEMS) and the Vaccine Impact on Diarrhea in Africa (VIDA) study, multisite case-control studies of moderate-to-severe diarrhea, the Malnutrition and Enteric Diseases (MAL-ED) study, a prospective observational birth cohort, the AntiBiotics for Children with Diarrhea (ABCD) trial, a multicenter placebo-controlled clinical trial of azithromycin for watery diarrhea, and the Enterics For Global Health (EFGH) – *Shigella* surveillance study (EFGH), a hybrid surveillance study of medically-attended diarrhea. Details of each study are included in the Supplementary Materials.

### Harmonization of covariates

Fecal samples were tested for enteric pathogens by quantitative PCR using the TaqMan Array Card (TAC) platform.^14^ Protocol details are included in the Supplementary Materials.

Moderate-to-severe diarrhea was defined to align with the enrollment criteria definition from GEMS and VIDA: any dehydration, dysentery (visible blood in stool), or hospitalization. Less severe diarrhea episodes were all those that did not meet this definition. Antibiotic treatment of each diarrhea episode was determined by record of receipt or prescription at enrollment (excluding antibiotics reported prior to diarrhea presentation) for the facility-based studies or caregiver-report during diarrhea at twice-weekly home visits for MAL-ED, as previously described.^15^ Antibiotic use was categorized into three groups: guideline recommended, possibly effective, and no or ineffective treatment. Guideline-recommended antibiotics included ciprofloxacin, azithromycin, ceftriaxone, and pivmecillinam.^16^ In MAL-ED, for which only drug classes were available, fluoroquinolones and macrolides were considered guideline-recommended. Antibiotics considered possibly effective were identified based on intrinsic activity with input from subject matter experts, and all other antibiotic use, including no treatment, was grouped into the “no or ineffective” category (Table S1). Children receiving multiple antibiotics were included in the most effective category of the antibiotics they received. For primary analyses, possibly effective antibiotics were collapsed with no or ineffective treatment into a single “no guideline recommended antibiotics” category given the similarity in results between those two groups.

The linear growth outcome was specified as length or height-for-age z-score (HAZ) as determined by WHO growth standards^17^ measured 60-90 days after the diarrhea episode depending on availability in the parent study. The HAZ measurement closest to 90 days following the episode (within ±45 days) was used in MAL-ED.

Common causes of *Shigella* and child growth as well as of antibiotic treatment and child growth were identified as confounding variables by causal diagram. Covariates representing these variables were selected based on data availability across studies and consistency with previous analyses. Baseline covariates included sex, age at enrollment in months, study site, socioeconomic status (SES) quintile by site, HAZ at the time of diarrhea (or the closest prior measurement within 75 days in MAL-ED), access to improved water, access to improved sanitation, number of children in the household under five years old, primary caregiver education, and days between diarrhea and follow-up measurement with 60-90 days. Episode severity characteristics included dysentery, fever, vomiting, dehydration level, maximum number of loose stools in 24 hours, and duration of episode before antibiotics. Additionally, we adjusted for log-transformed quantities of other enteric pathogens including rotavirus, adenovirus, ETEC, *Cryptosporidium*, astrovirus, norovirus GII, typical EPEC, *Campylobacter*, sapovirus, *Giardia*, *E. bieneusi*, and EAEC. Missing observations (∼1%, Table 1) were imputed using the study site-specific mean. Additional details of covariates are included in the Supplementary Materials.

**Table 1.**
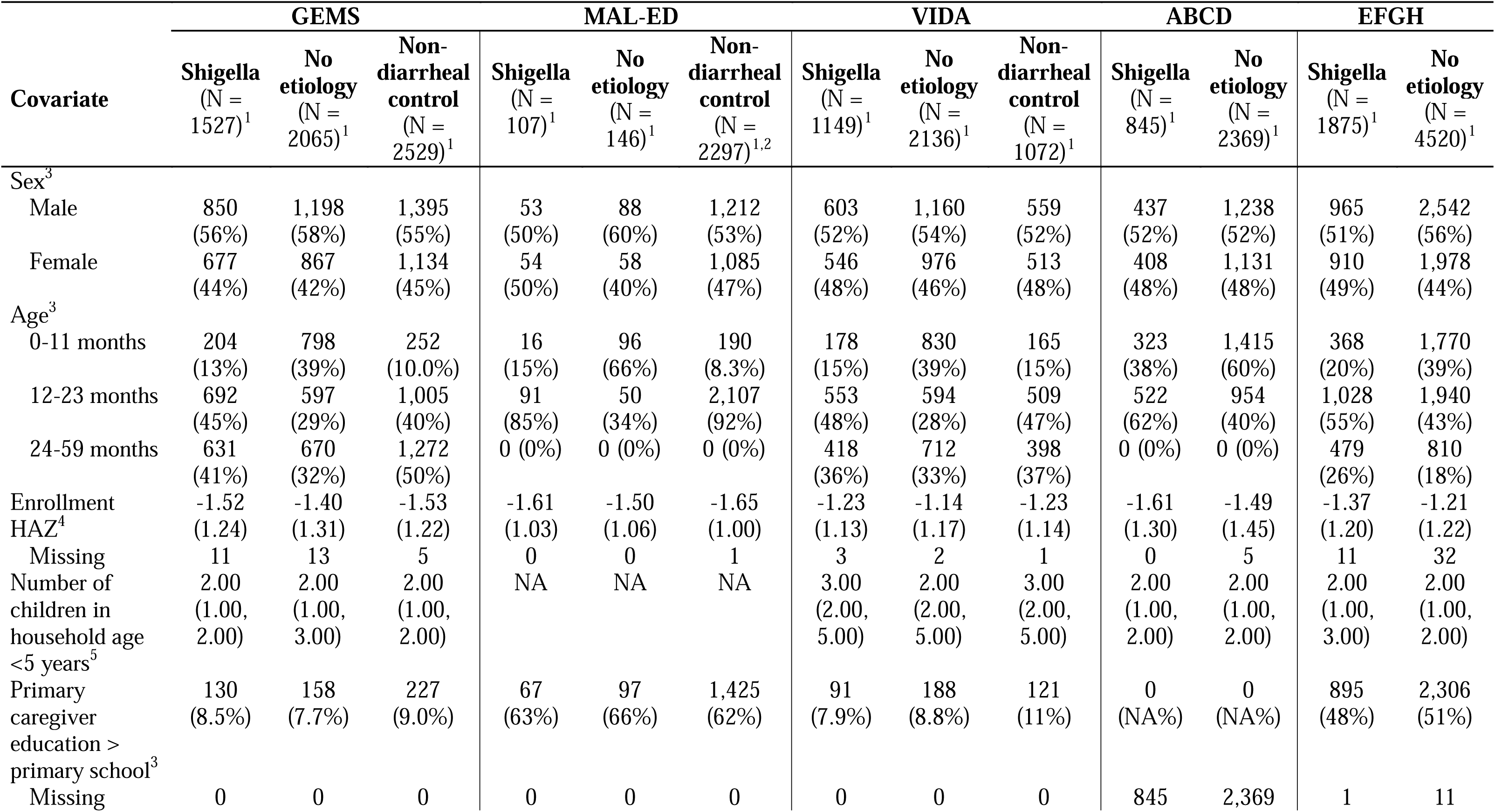
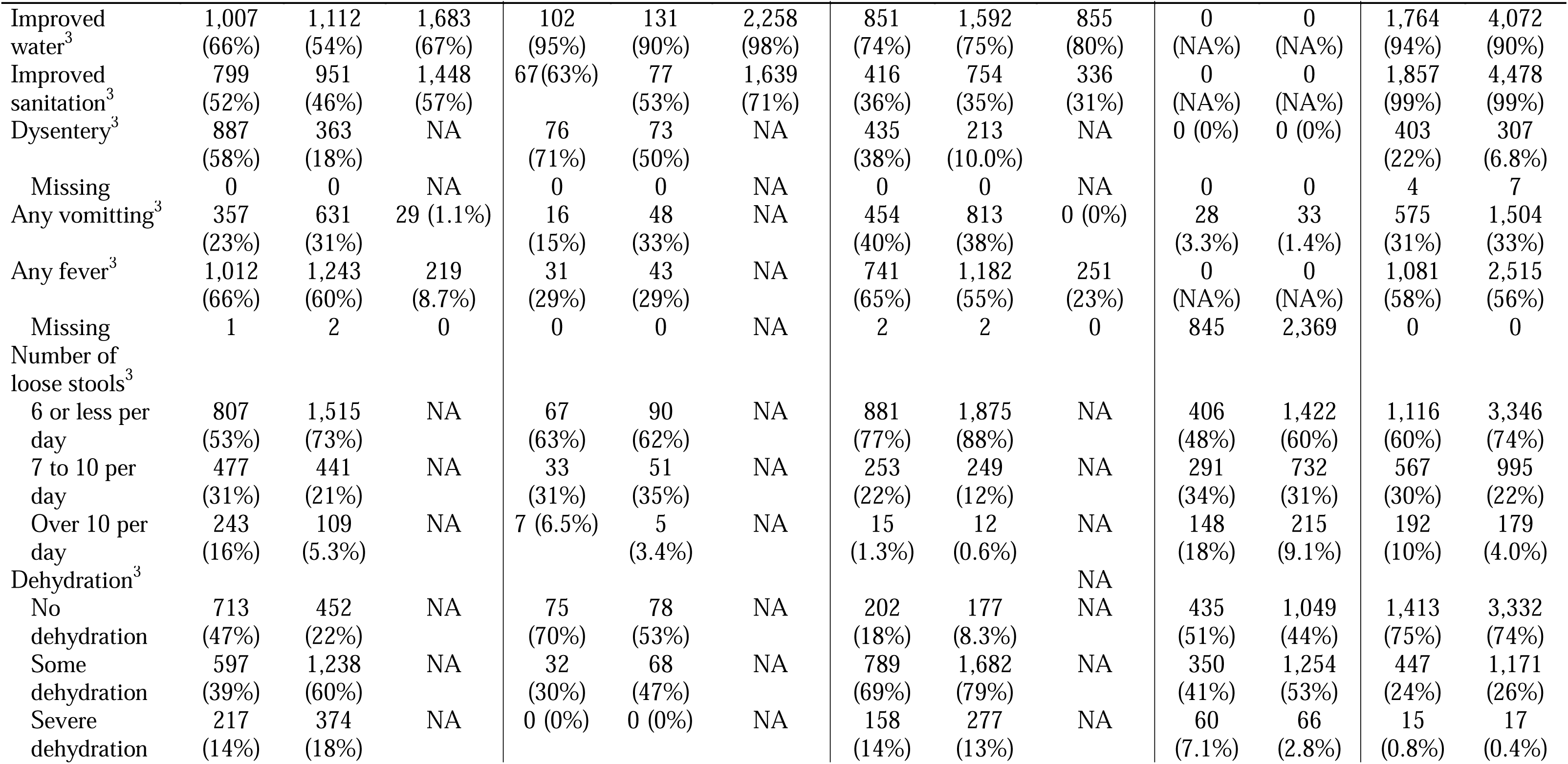
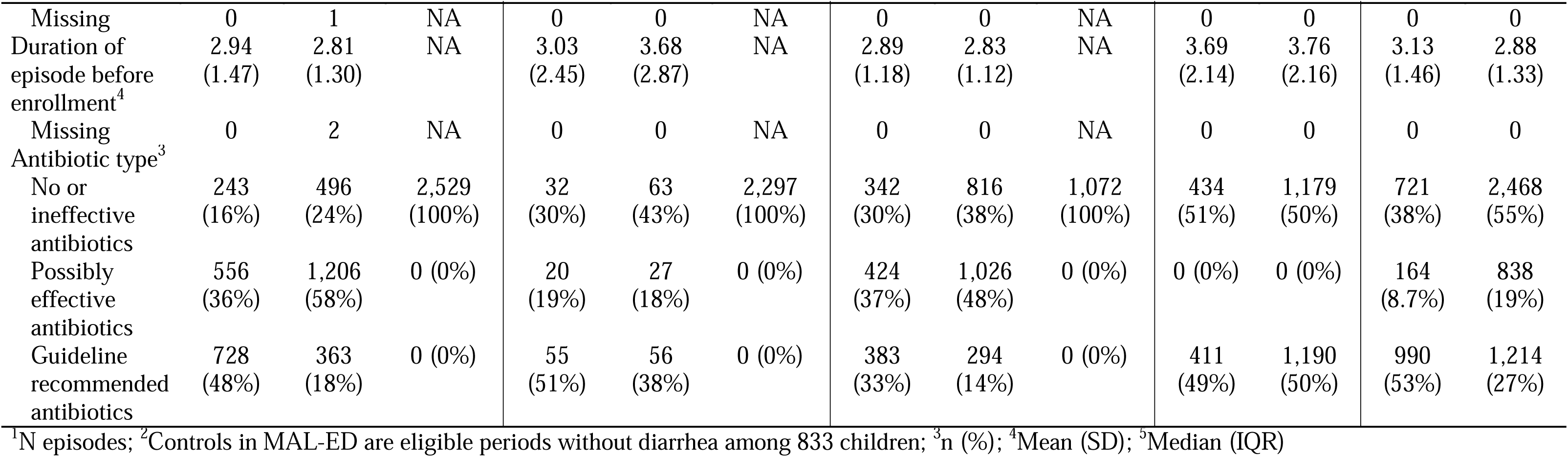
Characteristics of *Shigella*-attributable diarrhea episodes, episodes with no pathogen etiology identified, and non-diarrheal controls among the five studies included in the meta-analysis.

| Covariate | GEMS |  |  | MAL-ED |  |  | VIDA |  |  | ABCD |  | EFGH |  |
| --- | --- | --- | --- | --- | --- | --- | --- | --- | --- | --- | --- | --- | --- |
|  | Shigella<br>(N =<br>1527)<sup>1</sup> | No<br>etiology<br>(N =<br>2065)<sup>1</sup> | Non-diarrheal<br>control<br>(N =<br>2529)<sup>1</sup> | Shigella<br>(N =<br>107)<sup>1</sup> | No<br>etiology<br>(N =<br>146)<sup>1</sup> | Non-diarrheal<br>control<br>(N =<br>2297)<sup>1,2</sup> | Shigella<br>(N =<br>1149)<sup>1</sup> | No<br>etiology<br>(N =<br>2136)<sup>1</sup> | Non-diarrheal<br>control<br>(N =<br>1072)<sup>1</sup> | Shigella<br>(N =<br>845)<sup>1</sup> | No<br>etiology<br>(N =<br>2369)<sup>1</sup> | Shigella<br>(N =<br>1875)<sup>1</sup> | No<br>etiology<br>(N =<br>4520)<sup>1</sup> |
| Sex<sup>3</sup> |  |  |  |  |  |  |  |  |  |  |  |  |  |
| Male | 850<br>(56%) | 1,198<br>(58%) | 1,395<br>(55%) | 53<br>(50%) | 88<br>(60%) | 1,212<br>(53%) | 603<br>(52%) | 1,160<br>(54%) | 559<br>(52%) | 437<br>(52%) | 1,238<br>(52%) | 965<br>(51%) | 2,542<br>(56%) |
| Female | 677<br>(44%) | 867<br>(42%) | 1,134<br>(45%) | 54<br>(50%) | 58<br>(40%) | 1,085<br>(47%) | 546<br>(48%) | 976<br>(46%) | 513<br>(48%) | 408<br>(48%) | 1,131<br>(48%) | 910<br>(49%) | 1,978<br>(44%) |
| Age<sup>3</sup> |  |  |  |  |  |  |  |  |  |  |  |  |  |
| 0-11 months | 204<br>(13%) | 798<br>(39%) | 252<br>(10.0%) | 16<br>(15%) | 96<br>(66%) | 190<br>(8.3%) | 178<br>(15%) | 830<br>(39%) | 165<br>(15%) | 323<br>(38%) | 1,415<br>(60%) | 368<br>(20%) | 1,770<br>(39%) |
| 12-23 months | 692<br>(45%) | 597<br>(29%) | 1,005<br>(40%) | 91<br>(85%) | 50<br>(34%) | 2,107<br>(92%) | 553<br>(48%) | 594<br>(28%) | 509<br>(47%) | 522<br>(62%) | 954<br>(40%) | 1,028<br>(55%) | 1,940<br>(43%) |
| 24-59 months | 631<br>(41%) | 670<br>(32%) | 1,272<br>(50%) | 0 (0%) | 0 (0%) | 0 (0%) | 418<br>(36%) | 712<br>(33%) | 398<br>(37%) | 0 (0%) | 0 (0%) | 479<br>(26%) | 810<br>(18%) |
| Enrollment<br>HAZ<sup>4</sup> | -1.52<br>(1.24) | -1.40<br>(1.31) | -1.53<br>(1.22) | -1.61<br>(1.03) | -1.50<br>(1.06) | -1.65<br>(1.00) | -1.23<br>(1.13) | -1.14<br>(1.17) | -1.23<br>(1.14) | -1.61<br>(1.30) | -1.49<br>(1.45) | -1.37<br>(1.20) | -1.21<br>(1.22) |
| Missing | 11 | 13 | 5 | 0 | 0 | 1 | 3 | 2 | 1 | 0 | 5 | 11 | 32 |
| Number of<br>children in<br>household age<br><5 years<sup>5</sup> | 2.00<br>(1.00,<br>2.00) | 2.00<br>(1.00,<br>3.00) | 2.00<br>(1.00,<br>2.00) | NA | NA | NA | 3.00<br>(2.00,<br>5.00) | 2.00<br>(2.00,<br>5.00) | 3.00<br>(2.00,<br>5.00) | 2.00<br>(1.00,<br>2.00) | 2.00<br>(1.00,<br>2.00) | 2.00<br>(1.00,<br>3.00) | 2.00<br>(1.00,<br>2.00) |
| Primary<br>caregiver<br>education ><br>primary school<sup>3</sup> | 130<br>(8.5%) | 158<br>(7.7%) | 227<br>(9.0%) | 67<br>(63%) | 97<br>(66%) | 1,425<br>(62%) | 91<br>(7.9%) | 188<br>(8.8%) | 121<br>(11%) | 0<br>(NA%) | 0<br>(NA%) | 895<br>(48%) | 2,306<br>(51%) |
| Missing | 0 | 0 | 0 | 0 | 0 | 0 | 0 | 0 | 0 | 845 | 2,369 | 1 | 11 |
|  | Shigella<br>(N =<br>1527)<sup>1</sup> | No<br>etiology<br>(N =<br>2065)<sup>1</sup> | Non-diarrheal<br>control<br>(N =<br>2529)<sup>1</sup> | Shigella<br>(N =<br>107)<sup>1</sup> | No<br>etiology<br>(N =<br>146)<sup>1</sup> | Non-diarrheal<br>control<br>(N =<br>2297)<sup>1,2</sup> | Shigella<br>(N =<br>1149)<sup>1</sup> | No<br>etiology<br>(N =<br>2136)<sup>1</sup> | Non-diarrheal<br>control<br>(N =<br>1072)<sup>1</sup> | Shigella<br>(N =<br>845)<sup>1</sup> | No<br>etiology<br>(N =<br>2369)<sup>1</sup> | Shigella<br>(N =<br>1875)<sup>1</sup> | No<br>etiology<br>(N =<br>4520)<sup>1</sup> |
| Improved water<sup>3</sup> | 1,007<br>(66%) | 1,112<br>(54%) | 1,683<br>(67%) | 102<br>(95%) | 131<br>(90%) | 2,258<br>(98%) | 851<br>(74%) | 1,592<br>(75%) | 855<br>(80%) | 0<br>(NA%) | 0<br>(NA%) | 1,764<br>(94%) | 4,072<br>(90%) |
| Improved sanitation<sup>3</sup> | 799<br>(52%) | 951<br>(46%) | 1,448<br>(57%) | 67(63%) | 77<br>(53%) | 1,639<br>(71%) | 416<br>(36%) | 754<br>(35%) | 336<br>(31%) | 0<br>(NA%) | 0<br>(NA%) | 1,857<br>(99%) | 4,478<br>(99%) |
| Dysentery<sup>3</sup> | 887<br>(58%) | 363<br>(18%) | NA | 76<br>(71%) | 73<br>(50%) | NA | 435<br>(38%) | 213<br>(10.0%) | NA | 0 (0%) | 0 (0%) | 403<br>(22%) | 307<br>(6.8%) |
| Missing | 0 | 0 | NA | 0 | 0 | NA | 0 | 0 | NA | 0 | 0 | 4 | 7 |
| Any vomiting<sup>3</sup> | 357<br>(23%) | 631<br>(31%) | 29 (1.1%) | 16<br>(15%) | 48<br>(33%) | NA | 454<br>(40%) | 813<br>(38%) | 0 (0%) | 28<br>(3.3%) | 33<br>(1.4%) | 575<br>(31%) | 1,504<br>(33%) |
| Any fever<sup>3</sup> | 1,012<br>(66%) | 1,243<br>(60%) | 219<br>(8.7%) | 31<br>(29%) | 43<br>(29%) | NA | 741<br>(65%) | 1,182<br>(55%) | 251<br>(23%) | 0<br>(NA%) | 0<br>(NA%) | 1,081<br>(58%) | 2,515<br>(56%) |
| Missing | 1 | 2 | 0 | 0 | 0 | NA | 2 | 2 | 0 | 845 | 2,369 | 0 | 0 |
| Number of loose stools<sup>3</sup> |  |  |  |  |  |  |  |  |  |  |  |  |  |
| 6 or less per day | 807<br>(53%) | 1,515<br>(73%) | NA | 67<br>(63%) | 90<br>(62%) | NA | 881<br>(77%) | 1,875<br>(88%) | NA | 406<br>(48%) | 1,422<br>(60%) | 1,116<br>(60%) | 3,346<br>(74%) |
| 7 to 10 per day | 477<br>(31%) | 441<br>(21%) | NA | 33<br>(31%) | 51<br>(35%) | NA | 253<br>(22%) | 249<br>(12%) | NA | 291<br>(34%) | 732<br>(31%) | 567<br>(30%) | 995<br>(22%) |
| Over 10 per day | 243<br>(16%) | 109<br>(5.3%) | NA | 7 (6.5%) | 5<br>(3.4%) | NA | 15<br>(1.3%) | 12<br>(0.6%) | NA | 148<br>(18%) | 215<br>(9.1%) | 192<br>(10%) | 179<br>(4.0%) |
| Dehydration<sup>3</sup> |  |  |  |  |  |  |  |  | NA |  |  |  |  |
| No dehydration | 713<br>(47%) | 452<br>(22%) | NA | 75<br>(70%) | 78<br>(53%) | NA | 202<br>(18%) | 177<br>(8.3%) | NA | 435<br>(51%) | 1,049<br>(44%) | 1,413<br>(75%) | 3,332<br>(74%) |
| Some dehydration | 597<br>(39%) | 1,238<br>(60%) | NA | 32<br>(30%) | 68<br>(47%) | NA | 789<br>(69%) | 1,682<br>(79%) | NA | 350<br>(41%) | 1,254<br>(53%) | 447<br>(24%) | 1,171<br>(26%) |
| Severe dehydration | 217<br>(14%) | 374<br>(18%) | NA | 0 (0%) | 0 (0%) | NA | 158<br>(14%) | 277<br>(13%) | NA | 60<br>(7.1%) | 66<br>(2.8%) | 15<br>(0.8%) | 17<br>(0.4%) |
|  | Shigella<br>(N =<br>1527)<sup>1</sup> | No<br>etiology<br>(N =<br>2065)<sup>1</sup> | Non-diarrheal<br>control<br>(N =<br>2529)<sup>1</sup> | Shigella<br>(N =<br>107)<sup>1</sup> | No<br>etiology<br>(N =<br>146)<sup>1</sup> | Non-diarrheal<br>control<br>(N =<br>2297)<sup>1,2</sup> | Shigella<br>(N =<br>1149)<sup>1</sup> | No<br>etiology<br>(N =<br>2136)<sup>1</sup> | Non-diarrheal<br>control<br>(N =<br>1072)<sup>1</sup> | Shigella<br>(N =<br>845)<sup>1</sup> | No<br>etiology<br>(N =<br>2369)<sup>1</sup> | Shigella<br>(N =<br>1875)<sup>1</sup> | No<br>etiology<br>(N =<br>4520)<sup>1</sup> |
| Missing | 0 | 1 | NA | 0 | 0 | NA | 0 | 0 | NA | 0 | 0 | 0 | 0 |
| Duration of<br>episode before<br>enrollment<sup>4</sup> | 2.94<br>(1.47) | 2.81<br>(1.30) | NA | 3.03<br>(2.45) | 3.68<br>(2.87) | NA | 2.89<br>(1.18) | 2.83<br>(1.12) | NA | 3.69<br>(2.14) | 3.76<br>(2.16) | 3.13<br>(1.46) | 2.88<br>(1.33) |
| Missing | 0 | 2 | NA | 0 | 0 | NA | 0 | 0 | NA | 0 | 0 | 0 | 0 |
| Antibiotic type<sup>3</sup> |  |  |  |  |  |  |  |  |  |  |  |  |  |
| No or<br>ineffective<br>antibiotics | 243<br>(16%) | 496<br>(24%) | 2,529<br>(100%) | 32<br>(30%) | 63<br>(43%) | 2,297<br>(100%) | 342<br>(30%) | 816<br>(38%) | 1,072<br>(100%) | 434<br>(51%) | 1,179<br>(50%) | 721<br>(38%) | 2,468<br>(55%) |
| Possibly<br>effective<br>antibiotics | 556<br>(36%) | 1,206<br>(58%) | 0 (0%) | 20<br>(19%) | 27<br>(18%) | 0 (0%) | 424<br>(37%) | 1,026<br>(48%) | 0 (0%) | 0 (0%) | 0 (0%) | 164<br>(8.7%) | 838<br>(19%) |
| Guideline<br>recommended<br>antibiotics | 728<br>(48%) | 363<br>(18%) | 0 (0%) | 55<br>(51%) | 56<br>(38%) | 0 (0%) | 383<br>(33%) | 294<br>(14%) | 0 (0%) | 411<br>(49%) | 1,190<br>(50%) | 990<br>(53%) | 1,214<br>(27%) |

<sup>1</sup>N episodes; <sup>2</sup>Controls in MAL-ED are eligible periods without diarrhea among 833 children; <sup>3</sup>n (%); <sup>4</sup>Mean (SD); <sup>5</sup>Median (IQR)

### Statistical analysis

We defined *Shigella* cases as diarrhea episodes that were attributable to *Shigella* based on qPCR quantity and/or were positive by *Shigella* culture, among those with available qPCR data. In GEMS^1^ and MAL-ED,^2^ we used previously published attributable fractions (AFes) to determine attribution, classifying episodes as *Shigella* attributable if the AFe was greater than 0.5, as previously.^12^ Since the remaining studies did not have AFe data available, we instead used study-specific Ct cutoffs as defined in the original analyses for ABCD^14^ and EFGH.^18^ For VIDA, we used previously published Ct cut-offs from GEMS^1^ given the comparable sites and study design. *Shigella* culture data were unavailable for ABCD. To improve comparability of MAL-ED, the only study that identified diarrhea of any severity at home visits, we only included moderate-to-severe episodes from that study in the primary analysis.

We defined episodes of no known etiology as episodes for which qPCR results were available, but no pathogen tested met the AFe or Ct threshold, as appropriate per study, to be considered the cause of diarrhea. For comparison to non-diarrheal controls, matched controls were available for each diarrhea episode in GEMS and VIDA, matched to the index case on age, sex, site, enrollment time, and absence of diarrhea in the previous seven days. In MAL-ED, we applied similar criteria to match each diarrhea episode to all eligible children (1:2-1:58; median=1:20) without diarrhea in the previous seven days. The criteria were matched sex, site, and age (±2 months for episodes age 0-11 months, ±4 months for cases age 12-23 months), with a non-diarrheal sample collected within 15 days of the diarrhea sample.

Both analyses used augmented inverse probability weighted (AIPW) estimators to estimate controlled direct effects of *Shigella* on growth outcomes.^19^ This analysis treats antibiotic receipt as a mediator of the effect of *Shigella* on growth and accounts for potential time-varying confounding by adjusting for diarrheal severity indicators.^19^ In brief, this doubly robust approach flexibly modeled the growth outcome, and propensities for *Shigella* infection, antibiotic use, and outcome missingness using SuperLearner ensemble machine learning. Bias-corrected estimates of the controlled direct effect of *Shigella* on growth were then estimated by combining predictions from these models. We further used marginal structural models to assess effect heterogeneity by age. Additional details on the AIPW estimator, the machine learning models, and the marginal structural models are included in the Supplementary Materials.

We stratified analyses by age group, dysentery vs. watery diarrhea, and *Shigella* species (*S. flexneri* vs. *S. sonnei*). We also estimated the total effects of *Shigella* compared to non-diarrheal controls regardless of antibiotic use—overall, by diarrhea severity, and by age. We conducted several sensitivity analyses, including defining *Shigella* cases as culture-positive only and comparing *Shigella* cases to all other diarrhea episodes rather than only those with unknown etiology. In MAL-ED, since anthropometric measurements were collected monthly, we evaluated the total effects of *Shigella* on HAZ at each month, from one to 12 months following the episode, by diarrhea severity. Finally, in EFGH, we stratified effects on growth outcomes by antibiotic resistance among *Shigella* isolates from culture-positive episodes.

## Results

The meta-analysis comprised a total of 29,417 children, including 26,752 diarrhea episodes and 4,384 non-diarrheal controls who contributed 5,898 control periods. *Shigella* was attributed as the cause of diarrhea for 5,503 episodes. No etiology was identified for 11,236 episodes. Sociodemographic characteristics of diarrhea cases were largely similar across studies (Table 1). Children in MAL-ED and ABCD were younger than those in the other studies given the age limits on participation below 24 months. Children in GEMS and VIDA were more likely to present with dysentery, consistent with the moderate-to-severe diarrhea enrollment criteria in those studies. Across studies, children with *Shigella* diarrhea had lower enrollment HAZ than other children with diarrhea, and *Shigella* diarrhea episodes were more severe than episodes with no etiology identified (Table 1). Other enteric pathogens were more frequently detected in children with *Shigella*-attributed diarrhea compared to both episodes with no etiology identified and non-diarrheal controls (Table S2). Guideline recommended antibiotics were received for 46.6% of *Shigella* episodes, and antibiotics that may be effective against *Shigella* were received for 21.1% of *Shigella* episodes (Table 1). Guideline recommended treatment was most common for *Shigella* diarrhea episodes in EFGH (n=990, 53%) and least common in VIDA (n=383, 33%).

Compared to diarrhea episodes with no etiology identified, *Shigella*-attributable diarrhea without treatment with guideline recommended antibiotics was associated with small reductions in HAZ in the following 60-90 days in the IPD meta-analysis (HAZ difference: -0.03, 95% CI: -0.05, -0.01; Figure 1). Effects were similar for *Shigella* treated with possibly effective antibiotics (HAZ difference: -0.04, 95% CI: -0.06, -0.02) and treated with ineffective or no antibiotics (HAZ difference: -0.02, 95% CI: -0.04, -0.00; Figure S1). There was no effect of *Shigella* diarrhea on HAZ when treated with guideline recommended antibiotics (HAZ difference: -0.01, 95% CI: -0.02, 0.01). This pattern of effects was consistent in GEMS, ABCD, and EFGH, but not in VIDA and MAL-ED. In ABCD, the only study in which a guideline recommended antibiotic was randomized, the difference in the impact of treated and untreated *Shigella* was most pronounced, with placebo-treated episodes associated with a 0.07 (95% CI: 0.01, 0.13) decrement in HAZ compared to a 0.02 (95% CI: -0.04, 0.08) decrement for azithromycin treated episodes.

**Figure 1.**
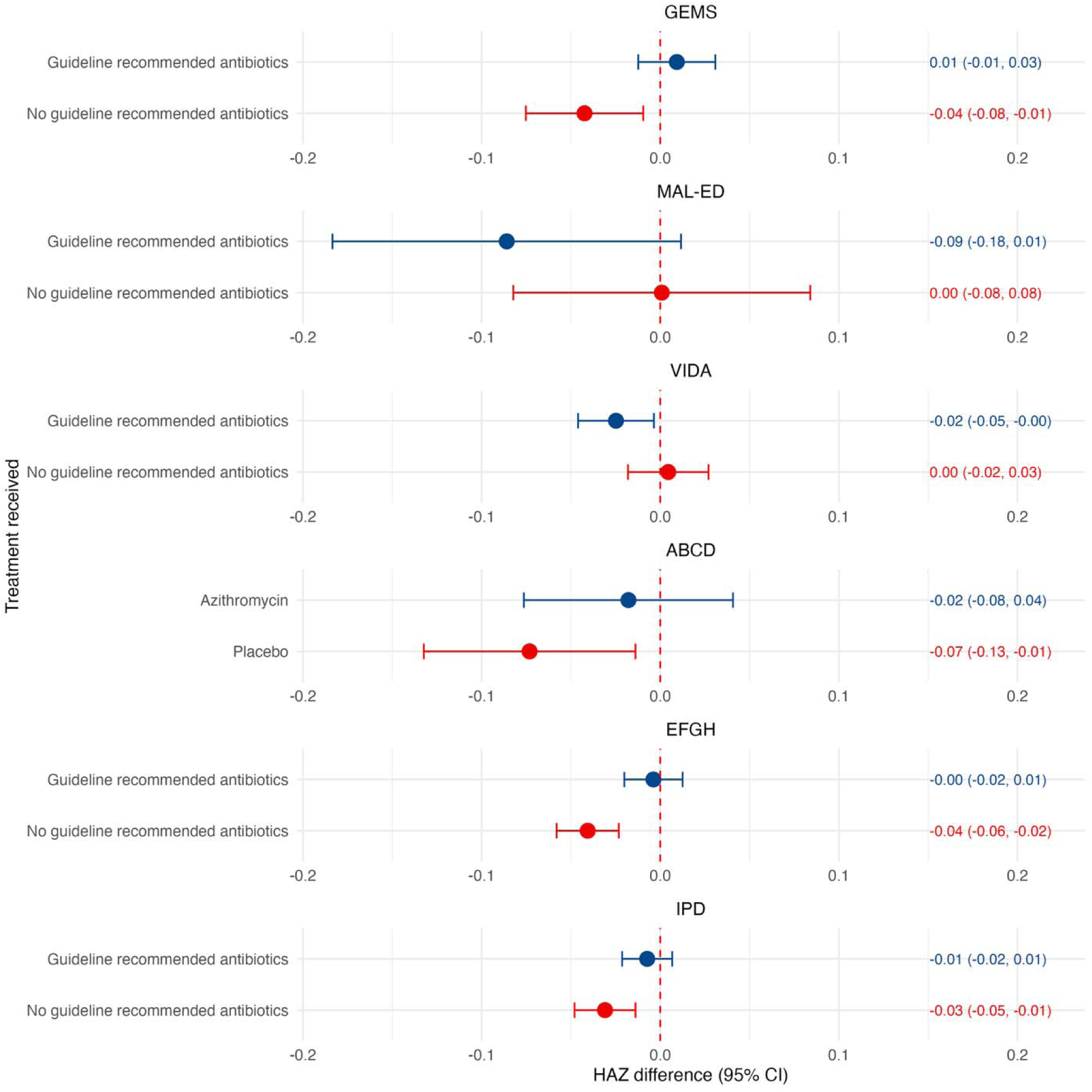
Effects of *Shigella*-attributable diarrhea on height-for-age z-score in the following 60-90 days compared to diarrhea with no pathogen etiology identified in each study and in an individual patient data (IPD) meta-analysis. Estimates are of the controlled direct effects of *Shigella* when treated with (blue) and without (red) guideline recommended antibiotics.

There was some evidence of heterogeneity in the effect of *Shigella* on HAZ by age overall (p for heterogeneity: 0.13, 0.03, 0.08 for episodes treated with guideline recommended antibiotics, possibly effective antibiotics, and no or ineffective antibiotics, respectively; Figure 2A). *Shigella* was associated with larger decrements in HAZ among younger children, regardless of antibiotics received, in all studies except GEMS (Figure S2).

**Figure 2.**
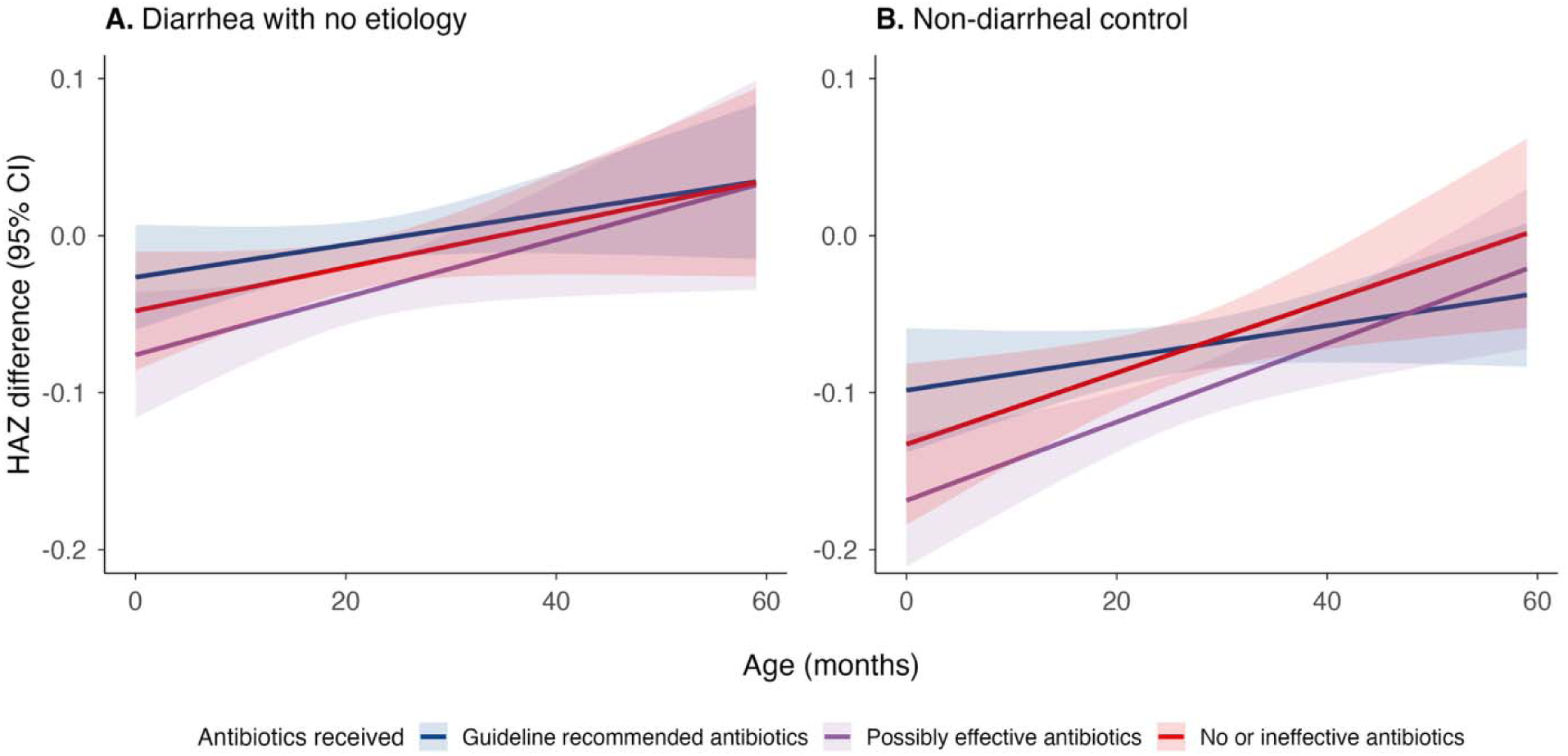
Heterogeneity by age in the effect of *Shigella*-attributable diarrhea on HAZ in the following 60-90 days in the IPD meta-analysis. **A.** compared to episodes with no pathogen etiology identified; **B.** compared to non-diarrheal controls.

Compared to non-diarrheal controls, *Shigella*-attributable diarrhea was associated with decrements in HAZ in the following 60-90 days regardless of antibiotic treatment (Figure S3). The overall effect was slightly larger for moderate-to-severe diarrhea episodes (HAZ difference: -0.09, 95% CI: -0.11, -0.07) than for less severe diarrhea episodes (HAZ difference: -0.07, 95% CI: -0.09, -0.04). Episodes that were treated with guideline recommended antibiotics had a slightly less pronounced negative effect on HAZ (difference: -0.08, 95% CI: -0.09, -0.06) than those that were not treated with guideline recommended antibiotics (HAZ difference: -0.11, 95% CI: -0.13, -0.09) (Figure 3), and this difference was driven by a larger negative effect in episodes treated with possibly effective antibiotics (Figure S4).

**Figure 3.**
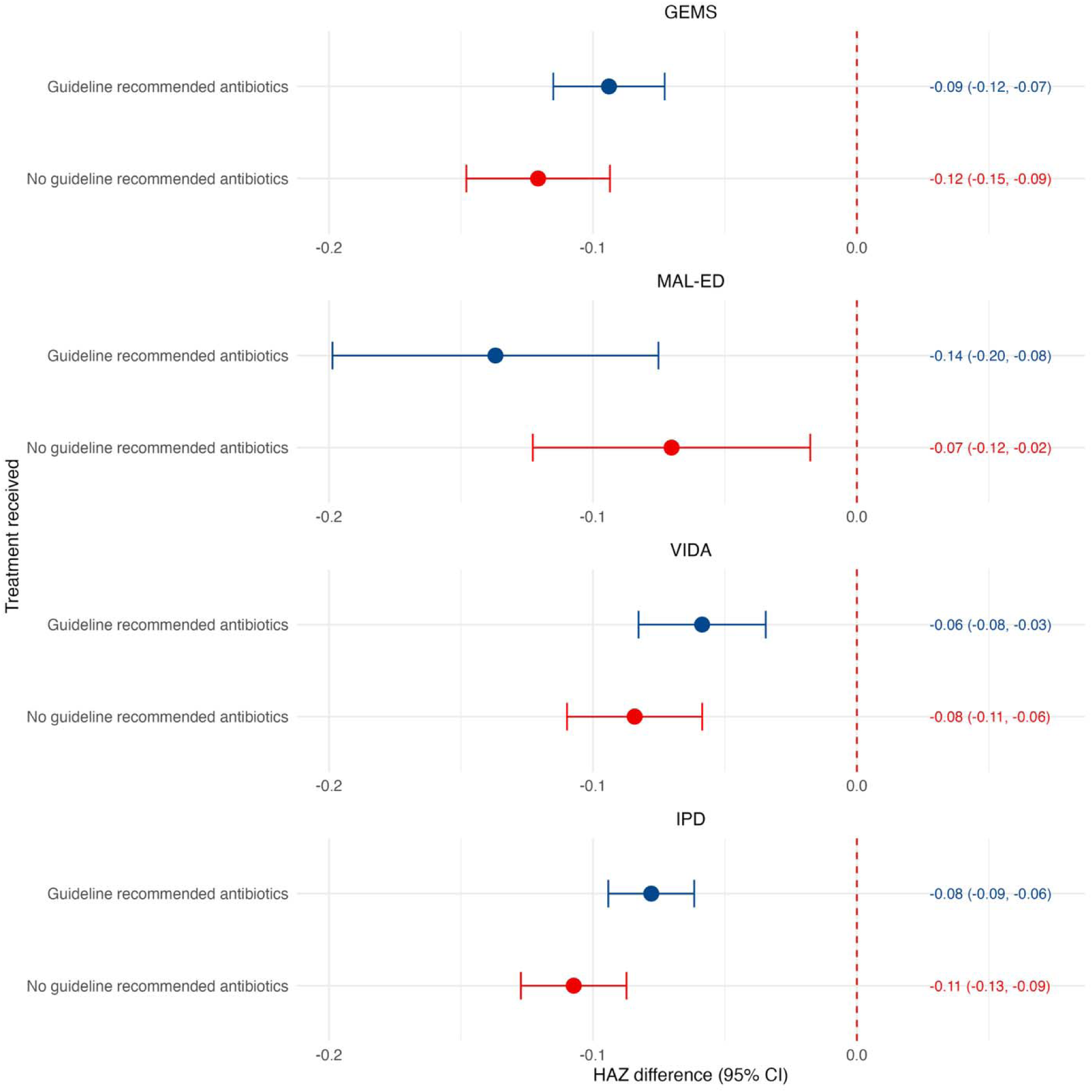
Effects of *Shigella*-attributable diarrhea on height-for-age z-score in the following 60-90 days compared to non-diarrheal controls in each study and in an individual patient data (IPD) meta-analysis. Estimates are of the controlled direct effects of *Shigella* when treated with (blue) and without (red) guideline recommended antibiotics.

The negative effect of *Shigella* on HAZ compared to non-diarrheal controls was larger among younger children (p for heterogeneity: 0.1, 0.001, 0.01 for episodes treated with guideline recommended antibiotics, possibly effective antibiotics, and no or ineffective antibiotics, respectively; Figure 2B). For example, *Shigella* among children 0-11 months of age was associated with a -0.10 decrement in HAZ (95% CI: -0.17, -0.03; Figure S5). This pattern of heterogeneity was not consistently observed in each study and antibiotic treatment group (Figure S6).

In MAL-ED, the magnitude of the effect of moderate-to-severe *Shigella* diarrhea increased from one to nine months following the episode (HAZ difference at 9 months: -0.24, 95% CI: -0.34, -0.15), after which there was catch-up growth (Figure 4). However, a growth decrement persisted at 12 months following the episode, of similar magnitude to that at three months. The effect was shorter lived for less-severe diarrhea, with the magnitude peaking four months after the episode and catch-up growth thereafter.

**Figure 4.**
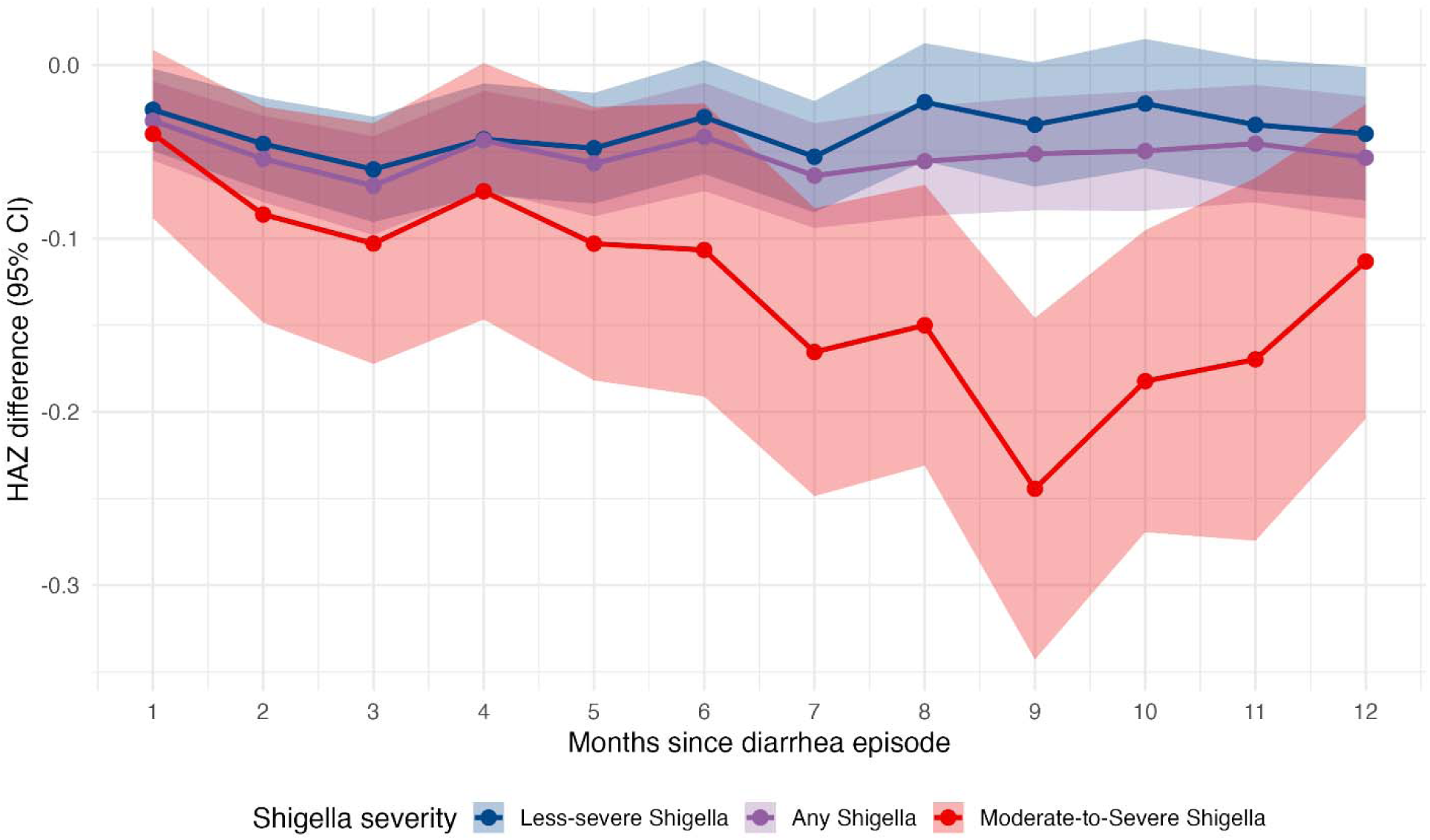
Effects of *Shigella*-attributable diarrhea, by diarrhea severity, on height-for-age z-score in the following 1-12 months compared to non-diarrheal controls in MAL-ED.

In an evaluation of whether antibiotic resistance among the isolated *Shigella* when treated with guideline recommended antibiotics modified the effect on HAZ in EFGH, *Shigella* episodes that were non-susceptible to the drug received were associated with larger decrements in HAZ (HAZ difference: -0.05, 95% CI: -0.07, -0.03) than *Shigella* episodes that were susceptible to the drug received (HAZ difference: -0.02, 95% CI: -0.04, 0.00) (Figure 5).

**Figure 5.**
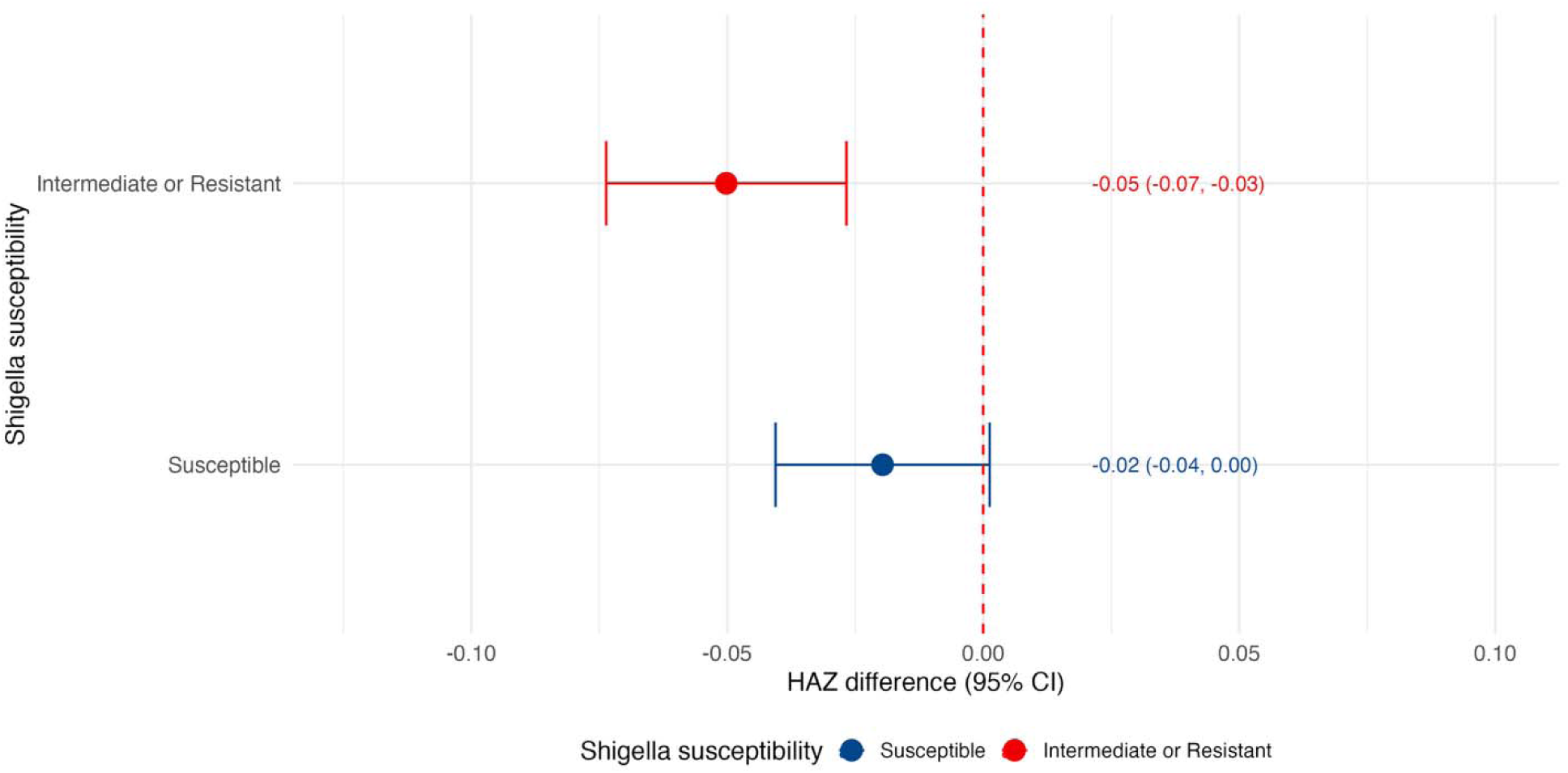
Among episodes treated with guideline recommended antibiotics in EFGH, controlled effects of *Shigella*-attributable diarrhea on height-for-age z-score in the following 90 days compared to diarrhea with no pathogen etiology, by *Shigella* antibiotic susceptibility. Estimates are of the controlled direct effects of *Shigella* when treated with an antibiotic the *Shigella* isolate was non-susceptible to (red), and *Shigella* when treated with an antibiotic the *Shigella* was susceptible to (blue).

With and without antibiotic treatment, the effects of *Shigella* dysentery were similar to those of *Shigella* watery diarrhea when compared to diarrhea episodes with no etiology identified (Figure S7). Compared to non-diarrheal controls, *Shigella* watery diarrhea had slightly larger effects than *Shigella* dysentery, though the age-stratified estimates were imprecise (Figure S8). *S. flexneri* and *S. sonnei* were associated with similar decrements in HAZ when compared to diarrhea episodes with no etiology and non-diarrheal controls, with slightly larger effects for *S. flexneri* under some treatments (Figure S9 and S10).

In a sensitivity analysis to evaluate the effect of culture-positive *Shigella* alone (excluding ABCD), the results were largely consistent (Figures S11 and S12). In a sensitivity analysis that compared *Shigella*-attributed diarrhea to all other observed diarrhea episodes, the effects of *Shigella* on HAZ were closer to the null in almost all cases and slightly positive in GEMS and VIDA under some treatments (Figure S13).

## Discussion

*Shigella*-attributed diarrhea was associated with small but significant decrements in HAZ in the short-term, with larger effects observed among younger children. Treatment with guideline-recommended antibiotics reduced the impact of *Shigella* compared to other diarrhea episodes but did not prevent effects on HAZ compared to non-diarrheal controls. The specific evidence that randomized azithromycin in ABCD prevented linear growth faltering associated with *Shigella* and that drug resistance lessened this protective effect in EFGH supports that the associations presented here quantify causal effects of *Shigella.* The findings suggest that a *Shigella* vaccine could improve growth among children for whom *Shigella* diarrhea is prevented. Appropriate antibiotic treatment may improve growth outcomes relative to other children with diarrhea but is unlikely to completely prevent decrements in linear growth.

The impact of *Shigella* on HAZ was largest among the youngest children. *Shigella* incidence increases steadily across the first 2 years of life and declines thereafter up to 5 years of age.^3,20^ While relatively lower incidence in the first year of life has been used to justify a later target vaccination schedule for *Shigella* vaccines,^5^ the evidence that *Shigella* has the largest impact on growth of children in the first year of life suggests a vaccine schedule completed early in infancy could be important to prevent *Shigella*-related growth faltering. Previous documentation of higher risk of death following *Shigella* among younger children^21^ further supports early administration of vaccine. The fact that there were no clear differences in the effect by disease phenotype (dysentery vs. watery diarrhea) highlights the important clinical impact of *Shigella* watery diarrhea and suggests expansions to treatment guidelines beyond dysentery may be warranted.

The effects estimated in the meta-analysis were largely consistent across included studies, and instances of heterogeneity may be explained by differences in study design and included diarrhea syndromes. ABCD was a randomized trial, such that confounding of the *Shigella*-treatment and treatment-growth relationships is unlikely. Therefore, it is not surprising that the difference in impact of *Shigella* when treated and untreated was most striking in this study. In the remaining studies, which were observational, residual confounding by indication (due to increased severity or other distinguishing characteristics of *Shigella* diarrhea relative to other episodes) may have biased estimates.

This harmonized analysis extends previously published analyses of the impact of *Shigella* diarrhea on linear growth faltering. In GEMS, untreated *Shigella* MSD was previously reported to be associated with a 0.16 z-score (95% CI: 0.04, 0.29) decrement in HAZ compared to other MSD episodes in the first year of life and a 0.06 z-score (95% CI: -0.01, 0.12) decrement in the second year.^11^ In MAL-ED, children with *Shigella* diarrhea compared to children with no diarrhea were reported to experience a 0.03 z-score (95% CI: 0.00, 0.05) decrement in LAZ in the following 90 days.^12^ Effects estimated in EFGH were very similar to those reported here.^13^ Other prior studies have also identified an association with *Shigella* and linear growth faltering with varying comparison groups and follow-up duration.^22,23^ For example, each *Shigella* diarrhea episode was associated with 0.08 cm decrement in linear growth in Peru,^24^ and *Shigella* diarrhea was significantly associated with change in length over one year in rural Bangladesh.^25^

However, these analyses either did not consider antibiotic treatment, or simply subset the analysis to episodes that were treated and untreated. Because antibiotic treatment is a mediator (rather than modifier) of the effect of *Shigella* on growth, this analytic approach may have led to bias. Specifically, children with *Shigella* may be more likely to be treated with antibiotics than children with other causes of diarrhea because of increased disease severity associated with *Shigella*. Specifically, *Shigella* causes dysentery which is a strong indication for antibiotic treatment. At the same time, disease severity may confound the association between antibiotic treatment and growth since severe disease may cause greater growth shortfalls. The present analysis accounts for both confounding by severity and the indirect effects of *Shigella* on growth through disease severity and antibiotic treatment.

Prior analyses were further complicated by choice of comparison group. In GEMS, *Shigella* diarrhea was compared to other diarrhea episodes, whereas in MAL-ED, children with *Shigella* diarrhea were compared to children without diarrhea. The latter comparison is more directly relevant to projections of vaccine impact since the vaccine would be expected to prevent *Shigella* diarrhea rather than convert it to diarrhea of another type, but this contrast cannot be estimated in studies without non-diarrheal controls. Furthermore, comparisons to other diarrhea episodes may not be generalizable since etiologies likely vary over time and geography. We considered two primary comparison groups: diarrhea episodes with no other etiology identified and non-diarrheal controls. Diarrhea episodes with no etiology identified are less affected by differences in the distribution of pathogens across sites and may be the closest proxy to a healthy child. In the sensitivity analysis in which we compared *Shigella* to all other diarrhea episodes, the associations with HAZ were smaller, which is as expected since other infectious causes of diarrhea have also been shown to be associated with poor growth.^11,12,26^

Because antibiotic resistance data were largely unavailable, we were unable to definitively classify whether an antibiotic received was effective for each episode. Treatment with possibly effective antibiotics did not reduce the impact of *Shigella* on HAZ, suggesting these drugs were in fact ineffective. When stratifying by whether antibiotics received were expected to be effective based on phenotypic resistance testing, as expected, *Shigella* had larger effects when they were non-susceptible to the antibiotics received.

While this meta-analysis was strengthened by identifying pathogen etiologies using quantitative molecular diagnostics, specific definitions for attribution varied slightly by study to account for differences in study design. For the facility-based observational studies, data on antibiotic treatment was limited by an inability to confirm adherence. The MAL-ED study collected daily information on antibiotic class use, but not specific drugs. Finally, our estimates may be conservative since the studies included involve sites with high engagement in research and recent socioeconomic status improvements, such that they may not be representative of more resource limited settings where *Shigella* impacts are greater.

In summary, this meta-analysis leveraged data spanning three continents and nearly two decades to estimate precise effects of *Shigella* on HAZ. The estimated effects of *Shigella* on HAZ are particularly informative for future modeling studies of vaccine impact on child growth, which are likely to continue to be of interest as *Shigella* vaccines advance through the development pipeline.

## Data Availability

All data included in the present study are available upon reasonable request to the authors. Data from GEMS and MAL-ED are available online at clinepidb.org. Data from EFGH are available at https://doi.org/10.25934/PR00011860.

https://clinepidb.org/ce/app/workspace/analyses/DS_5c41b87221/new/variables/PCO_0000024/ENVO_00000009

https://clinepidb.org/ce/app/workspace/analyses/DS_841a9f5259/new/variables/PCO_0000024/ENVO_00000009

https://doi.org/10.25934/PR00011860

## Data sharing

GEMS, MAL-ED, and EFGH data are publicly available. VIDA and ABCD data are available by request.

