## Supplemental Materials for "Effects of *Shigella* diarrhea with and without antibiotic treatment on linear growth: an individual patient data meta-analysis of five multisite studies among children in low-resource settings"

### Supplementary Methods

#### Details of included studies

*GEMS*

The Global Enteric Multicenter Study (GEMS) was a multisite case-control study of moderate-to-severe diarrhea in seven sites in The Gambia, Kenya, Mali, Mozambique, Bangladesh, India and Pakistan from 2007-2011.[1] Children aged 0-59 months were enrolled as cases if they presented with an acute episode of moderate-to-severe diarrhea, defined as three or more loose stools within the last 24 hours, with sunken eyes, loss of skin turgor, intravenous hydration, dysentery, or admission to hospital. Controls, without diarrhea in the previous 7 days, were randomly matched by age, sex, and residence and enrolled within 14 days of the case. Anthropometric measurements and stool samples were taken at enrollment, and children were re-measured at a 60-day follow-up visit to assess vital status and interim medical events.[1]

*MAL-ED*

The Malnutrition and Enteric Diseases (MAL-ED) study was a prospective observational birth cohort conducted at 8 low-resource sites in Brazil, Peru, Tanzania, South Africa, Pakistan, India, Nepal, and Bangladesh.[2] From 2009-2014, enrolled children were followed twice-weekly for surveillance of illnesses and feeding practices at home visits until two years of age.[3] Weight and length were measured monthly, and stool samples were collected monthly and during diarrhea. Diarrhea was defined by maternal report of three or more loose stools in 24 hours or visible blood in stool.[4]

*VIDA*

The Vaccine Impact on Diarrhea in Africa (VIDA) study was a case-control study of moderate-to-severe diarrhea conducted from 2015-2018 in three African sites (The Gambia, Mali, and Kenya) that previously participated in GEMS.[5] The study design, data collection, and microbiological methods were designed to follow the GEMS protocols to allow comparison between studies.

*ABCD*

The AntiBiotics for Children with Diarrhea (ABCD) trial was a multicenter, double-blinded, randomized, parallel-group, placebo-controlled clinical trial conducted in seven sites in Bangladesh, India, Kenya, Malawi, Mali, Pakistan, and Tanzania.[6,7] Between 2017-2019, the trial enrolled children aged 2-23 months with acute watery diarrhea (3+ watery stools in the previous 24 hours). Eligible children had dehydration, moderate wasting (weight-for-length z-score between -2 and -3 or a mid-upper arm circumference between 115 mm and 125 mm), and/or severe stunting (length-for-age z-score < -3).[6] Children were excluded if they had dysentery, suspected cholera, severe acute malnutrition, other infections requiring antibiotics, or had received antibiotics in the last 14 days. Children were randomized 1:1 to receive a 3-day course of azithromycin (10 mg/kg) or placebo. At enrollment, sociodemographic (age, sex, socioeconomic status, household size) and diarrhea severity characteristics (vomiting, number of loose stools, diarrhea duration, dehydration) were recorded. Anthropometry (middle-upper arm circumference, weight, and length) and fecal samples were also collected. Follow-up visits ascertained total diarrhea duration, hospitalizations through day 90, anthropometry at day 90, and vital status.

*EFGH*

The Enterics For Global Health (EFGH) – *Shigella* surveillance study (EFGH) was a hybrid surveillance study of medically-attended diarrhea conducted from 2022-2024 in seven sites in Bangladesh, Kenya, Malawi, Mali, Pakistan, Peru, and The Gambia. Children aged 6-35 months presenting at medical facilities with diarrhea (≥3 abnormally loose or watery stools in the previous 24 hours with or without blood) within 7 days of onset were enrolled. At enrollment, sociodemographic and clinical characteristics were documented, anthropometry was measured, and fecal samples, including rectal swabs and whole stool if available, were collected. A diarrhea diary was used for 14 days to document diarrhea severity and medications. Follow-up visits at 4 weeks and 3 months included questionnaires to document recovery from diarrhea, new diarrhea episodes, hospitalizations, and deaths. Anthropometry was repeated at 3 months.

**Details of pathogen diagnostic methods**

Total nucleic acid was extracted using the QIAamp Stool Fast DNA Mini kit (Qiagen, Valencia, California), and qPCR was conducted using AgPath One Step reverse-transcription PCR reagents (Thermo Fisher, Carlsbad, California). External controls (MS2 for RNA targets and phocine herpesvirus for DNA targets) were spiked into each sample to monitor extraction and amplification performance. One extraction blank was included to monitor laboratory contamination. A cycle threshold (Ct) value of 35 was considered the analytical limit of detection, as previously.[8] Ct values from rectal swabs were adjusted by the mean Ct difference for each pathogen between paired rectal swabs and whole stools to account for differences in sensitivity between sample types.[8]

**Details of harmonization of covariates**

Socioeconomic status (SES) was incorporated into both the individual study and pooled IPD analyses using site-specific SES quintiles derived from available socioeconomic information. MAL-ED and EFGH provided the WAMI (water, assets, maternal education, and income) and SES index scores, respectively, which we converted to site-specific quintiles. GEMS and VIDA lacked summary scores, so we constructed principal-components analysis (PCA) scores from enrollment questionnaire variables and calculated quintiles within sites. ABCD directly reported a site-specific SES quintile variable, which was used as provided.

Analyses of individual studies used the most granular form of each covariate where available and omitted covariates that were unavailable in that study.

In the pooled IPD analysis, covariates were harmonized to align definitions as closely as possible across studies. Water and sanitation access were binarized in each study as improved versus unimproved. Similarly, primary caregiver education was binarized as greater than primary school versus primary school or less. ABCD did not collect either of these variables, so all observations were coded as unimproved for water and sanitation, and primary school or less for education. The number of children under age five years in the household was not collected in MAL-ED, so it was set to one for all observations, assuming no other young children in the household.

As with covariates, most of the severity information was harmonized for the pooled analysis by collapsing variables into binary or categorical formats that were uniform across studies. Vomiting and fever were coded as binary indicators for any occurrence. The maximum number of loose stools was categorized 6 or less, 7 to 10, and 10 or more. Dehydration was broken into none, some, or severe. ABCD did not collect information on fever, so this variable was coded as "no" for all observations. Dysentery was an exclusion criterion in ABCD, so all participants were coded as not having dysentery. Similarly, moderate to severe diarrhea (MSD) was an inclusion criterion for GEMS and VIDA. The GEMS definition of MSD was used to define MSD in MAL-ED.

Phenotypic serotyping data were available for culture-positive *Shigella* episodes in GEMS, VIDA, and EFGH. Genotypic serotyping data were available for qPCR-attributable *Shigella* in MAL-ED and EFGH. For the sensitivity analysis stratifying by *Shigella* species, we included all cases that had a species identified either by phenotypic or genotypic data wherever available. Serotyping data were not available in ABCD; thus, samples from ABCD were excluded from this analysis.

**Primary analysis methods**

We applied an augmented inverse probability weighted (AIPW) estimator to estimate the effect of *Shigella* and antibiotics on growth in the subset of children who acquired *Shigella* diarrhea.

This estimator combines predictions from regression models to build a two-stage estimate. In the first stage, a plug-in estimator is computed based on modeling HAZ as a function of *Shigella* diarrhea, antibiotic usage, and confounders. In the second stage, a bias correction is applied utilizing propensity scores that estimate the probability of *Shigella* diarrhea and antibiotic usage. We also estimate the propensity for missing outcome data to correct for potential informative missingness.

The final, bias-corrected AIPW estimate is multiply robust, meaning its estimates are approximately unbiased for the true effect if some combination of the regression estimates are consistent for their true values. Use of this method to estimate the causal effect allows us to appropriately account for the role of antibiotic treatment as a mediator of *Shigella* and growth, as well as the role of disease severity as a confounder of antibiotics and growth and mediator of *Shigella* and growth (see the directed acyclic graph; DAG below).

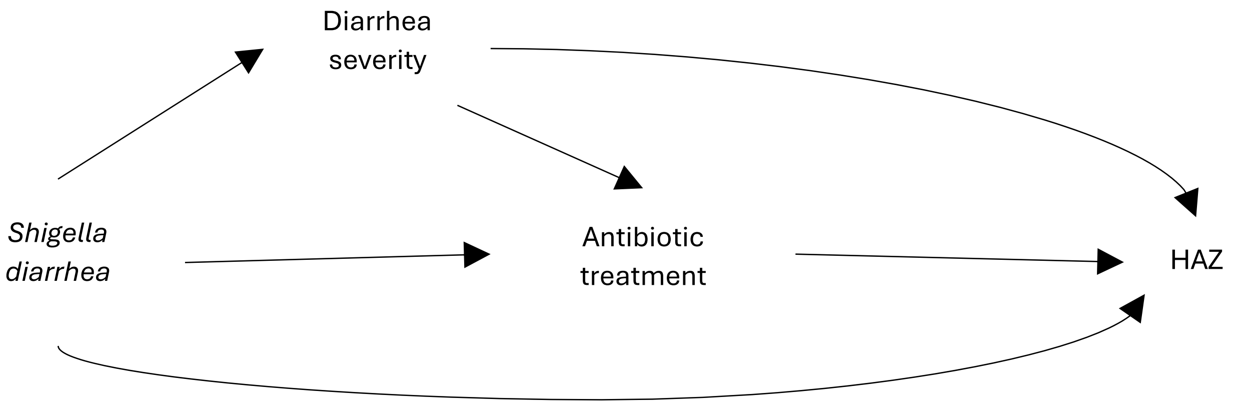


**Super Learning**

An additional benefit of the AIPW estimator is that it allows for the use of machine learning to flexibly model the nuisance regressions, adding an additional layer of robustness, while ensuring well-calibrated confidence intervals and hypothesis tests. We take advantage of this property by applying Super Learner, an ensemble modeling method, to estimate each of the regressions described below. Super Learner uses cross-validation to combine several candidate models into an optimally weighted ensemble model.[9,10] The final ensemble model has statistical properties guaranteeing its accuracy matches or exceeds that of its candidate models. By using Super Learner, we are able to explore a range of models for each nuisance regression and use a data-driven approach for regression estimator selection. The library of candidate regressions used in Super Learner was

1. Main terms generalized linear model (SL.glm) with a screening step taken for models that included pathogen quantities to eliminate pathogens detected in <5% of samples
2. Main terms generalized linear model (SL.glm) with a screening step to include variables with non-zero coefficients in a Lasso regression (screen.glmnet)
3. Generalized linear model with natural cubic splines for age and baseline HAZ with a screening step taken for models that included pathogen quantities to eliminate pathogens detected in <5% of samples.
4. Stepwise forward selection model with splines for age and baseline HAZ. For models that adjust for antibiotic use, antibiotics were forced to be included in the model.
5. Lasso regression (SL.glmnet)
6. Random forest (SL.ranger)
7. Multivariate adaptive regression splines (SL.earth)

Gradient boosted trees (SL.xgboost)

Super learning was used to estimate each of the following regressions.

**Outcome Regressions**

The AIPW approach requires fitting several regressions to model the linear growth outcome. These regressions were estimated separately in subsets of the data corresponding to *Shigella* attributable diarrhea, no-etiology diarrhea, and controls.

For the *Shigella* and no etiology diarrhea subsets, we regressed follow-up HAZ on antibiotic treatment, baseline covariates, severity indicators, and additional pathogen quantities. In the control subset, we regressed HAZ on baseline covariates only. After fitting each model in its respective subset, we evaluated the model fit under each level of antibiotic treatment on the full dataset.

In the no etiology diarrhea group, we fit a second stage regression model of the predictions from the first-stage outcome regression on baseline covariates only. This second stage model allows us to appropriately average the regression estimates over the distribution of diarrhea severity indicators in children with diarrhea with no attributable etiology. This marginalization ensures that differences in diarrhea severity due to no etiology vs. *Shigella* diarrhea are appropriately reflected in our effect estimates.

The final outcome model predictions were marginalized over individuals with *Shigella*-attributable diarrhea to obtain an initial, plug-in estimate of the effect of interest.

**Propensity Regressions**

Next, we estimated propensity for (i) antibiotic use, (ii) infection, and (iii) outcome missingness to perform a bias-correction of the effect estimates.

*Antibiotic Propensity*

To estimate the probability of receiving each level of antibiotics, we used a sequential modeling approach. For each antibiotic level, we estimated a regression function that regressed a binary indicator of receiving the given level of antibiotics on baseline covariates, severity indicators, and pathogen quantities, within the subset of participants who had not received antibiotics from the previously modeled levels. Models were fit separately for the *Shigella* diarrhea and no etiology subsets. These antibiotic use models were not used to estimate growth outcomes in controls.

To illustrate the sequential approach using our three-level scenario (guideline recommended, possibly effective, and no or ineffective antibiotics), we first modeled the probability of receiving guideline recommended antibiotics among all participants in the relevant infection-status subset. Next, we modeled the probability of receiving possibly effective antibiotics among those who did not receive guideline recommended antibiotics and multiplied it by one minus the probability of receiving guideline-recommended antibiotics to obtain estimates of the conditional probability of receiving possibly effective antibiotics. Lastly, we assigned the probability of receiving no or ineffective antibiotics as one minus the sum of the previous two probabilities. This ensures that the predicted probabilities across antibiotic levels sum to one for each individual.

*Infection Propensity*

The propensity model for *Shigella* infection was fit by regressing a binary indicator of *Shigella* attributable diarrhea on baseline covariates in the full dataset. The propensity model for diarrhea of no known etiology was fit by regressing a binary indicator of diarrhea with no known etiology on baseline covariates within the subset of people who did not have *Shigella* attributable diarrhea. Infection propensity models were not applicable for controls.

*Missingness of Follow-up HAZ Propensity*

To model the probability of missing follow-up HAZ, we fit separate propensity models within the *Shigella,* no etiology, and control subsets. In the *Shigella* and no etiology subsets, we regressed an indicator of missingness on antibiotic use, baseline covariates, severity indicators, and pathogen quantities. The propensity model in the control subset included baseline covariates only.

**AIPW Effect Estimation**

After fitting each propensity model and generating relevant predictions, we truncated extreme propensity scores to reduce instability.[11] We then combined predictions to construct the second-stage bias correction term. This was added to each initial plug-in estimate to produce the final AIPW estimate of expected growth given antibiotic use and Shigella infection for each level of antibiotic use and infection. Effect estimates were calculated by taking the difference between expected growth under *Shigella* versus no etiology, and *Shigella* versus control. Variance estimates were derived using estimates of the efficient influence functions. Additional adjustments were applied to account for participant re-enrollment in the study.[12]

**Effect heterogeneity by age**

Linear marginal structural models were utilized to summarize effect heterogeneity of *Shigella* diarrhea by age. We utilize AIPW estimators for the parameters of the marginal structural model, which can be derived via straightforward extensions of the methods described in Rosenblum and van der Laan (2010).[13] These estimators are built using the same outcome regression estimates and propensity scores that are used in the primary analysis. To test for effect heterogeneity, we performed Wald-style hypothesis tests of the slope parameter for age in the model. Standard errors for the test were computed based on estimates of the efficient influence function. Importantly, our hypothesis tests have a model-free interpretation as a test for trend in the magnitude of effects across ages.[14] In other words, the true effect modification could be non-linear and our results would still yield an interpretable test for trend.

**Antibiotic resistance sensitivity analysis**

To evaluate the impact of antibiotic resistance on growth outcomes, we conducted a sensitivity analysis using antimicrobial susceptibility testing data for culture positive samples in the EFGH study among episodes that received guideline recommended antibiotics. This analysis was restricted to the subset of participants with available TAC data to ensure consistency with the comparison group used in the primary analysis.

Episodes were considered antibiotic non-susceptible if the episode was treated with an antibiotic that the corresponding *Shigella* isolate was intermediate or resistant to by antimicrobial susceptibility testing using the Kirby-Bauer disc diffusion method.[15] Episodes were classified as susceptible if the isolate was susceptible to any of the guideline recommended antibiotics received during the episode. We applied the same AIPW and Super Learner methods as in the primary analysis to estimate expected growth outcomes. We contrasted estimates of (i) the effect of *Shigella* diarrhea when caused by *Shigella* that is susceptible to any guideline-recommended antibiotic administered versus diarrhea with no attribution treated with guideline-recommended antibiotics; and (ii) the effect of *Shigella* diarrhea when caused by *Shigella* that is intermediate or resistant to all guideline-recommended antibiotics administered versus diarrhea with no attribution treated with guideline-recommended antibiotics.

**Table S1.** Classification of antibiotics included in the analysis.

| Guideline recommended | Possibly effective | Ineffective |
|---|---|---|
| Ciprofloxacin | Amikacin | Biodroxil |
| Azithromycin | Amoxicillin | Cefadroxil |
| Ceftriaxone | Ampicillin | Cephalexin |
| Pivmecillinam | Ampiclox | Cephradine |
| | Augmentin | Clindamycin |
| | Cefaclor | Cloxacillin |
| | Cefixime | Entamizole |
| | Cefotaxime | Enteroquinole |
| | Cefpodoxime | Fluconazole |
| | Ceftazidime | Flucloxacillin |
| | Cefuroxime | Metronidazole |
| | Cephalosporins | Mupirocin |
| | Chloramphenicol/Thiamphenicol | Nitazoxamide |
| | Clarithromycin | Orex |
| | Doxycycline | Penicillin |
| | Erythromycin | Pyrazinamide |
| | Furazolidone | Spornidex |
| | Gentamycin | Tetracycline |
| | Levofloxacin | Unknown class or drug |
| | Meropenam | |
| | Nalidixic acid | |
| | Nifuroxazide | |
| | Nitrofurantoin | |
| | Norfloxacin | |
| | Ofloxacin | |
| | Streptomycin | |
| | Sulfamide | |
| | Sulfonamides | |
| | Thiamphenicol | |
| | Trimethoprim-sulfamethoxazole | |

**Table S2**. Pathogens detected by qPCR among *Shigella*-attributed diarrhea episodes, episodes with no etiology identified, and non-diarrheal controls.

| | **GEMS** | | | **MALED** | | | **VIDA** | | | **ABCD** | | **EFGH** | |
|---|---|---|---|---|---|---|---|---|---|---|---|---|---|
| **Pathogen detection by qPCR** | **Shigella** (N = 1527)[1] | **No etiology** (N = 2065)[1] | **Non-diarrheal control** (N = 2529)[1] | **Shigella** (N = 107)[1] | **No etiology** (N = 146)[1] | **Non-diarrheal control** (N = 2297)[12] | **Shigella** (N = 1149)[1] | **No etiology** (N = 2136)[1] | **Non-diarrheal control** (N = 1072)[1] | **Shigella** (N = 845)[1] | **No etiology** (N = 2369)[1] | **Shigella** (N = 1875)[1] | **No etiology** (N = 4520)[1] |
| *Shigella* detected | 1,509 (99%) | 424 (21%) | 432 (29%) | 107 (100%) | 13 (8.9%) | 416 (18%) | 1,131 (98%) | 301 (14%) | 365 (34%) | 845 (100%) | 187 (7.9%) | 1,828 (97%) | 574 (13%) |
| Missing | 0 | 0 | 1,064 | 0 | 0 | 19 | 0 | 0 | 0 | 0 | 0 | 0 | 0 |
| Rotavirus detected | 174 (12%) | 78 (3.8%) | 76 (5.2%) | 13 (12%) | 10 (6.8%) | 66 (2.9%) | 61 (5.3%) | 19 (0.9%) | 30 (2.8%) | 117 (14%) | 81 (3.4%) | 176 (9.5%) | 82 (1.9%) |
| Missing | 15 | 18 | 1,075 | 2 | 0 | 25 | 7 | 16 | 11 | 1 | 3 | 27 | 127 |
| Adenovirus detected | 435 (29%) | 429 (21%) | 338 (23%) | 36 (34%) | 44 (30%) | 451 (20%) | 171 (15%) | 158 (7.6%) | 149 (14%) | 190 (23%) | 271 (12%) | 384 (21%) | 446 (10%) |
| Missing | 20 | 15 | 1,072 | 1 | 0 | 30 | 33 | 46 | 26 | 13 | 47 | 84 | 125 |
| ETEC detected | 663 (44%) | 777 (38%) | 633 (43%) | 47 (45%) | 48 (33%) | 935 (41%) | 555 (49%) | 709 (33%) | 497 (46%) | 421 (50%) | 558 (24%) | 824 (45%) | 1,106 (25%) |
| Missing | 14 | 12 | 1,070 | 2 | 0 | 18 | 10 | 11 | 2 | 4 | 15 | 35 | 33 |
| *Cryptosporidium* detected | 296 (20%) | 350 (17%) | 202 (14%) | 9 (8.7%) | 13 (8.9%) | 273 (12%) | 260 (23%) | 352 (17%) | 230 (22%) | 205 (24%) | 269 (11%) | 271 (15%) | 377 (8.4%) |
| Missing | 10 | 6 | 1,065 | 3 | 0 | 23 | 26 | 20 | 13 | 0 | 4 | 11 | 28 |
| Astrovirus detected | 109 (7.2%) | 95 (4.6%) | 70 (4.8%) | 21 (20%) | 31 (21%) | 302 (13%) | 61 (5.4%) | 73 (3.4%) | 39 (3.7%) | 86 (10%) | 109 (4.6%) | 70 (3.8%) | 135 (3.0%) |
| Missing | 13 | 20 | 1,069 | 2 | 0 | 25 | 9 | 13 | 13 | 1 | 5 | 19 | 77 |
| Norovirus GII detected | 154 (10%) | 254 (12%) | 152 (10%) | 15 (14%) | 19 (13%) | 377 (17%) | 92 (8.1%) | 251 (12%) | 100 (9.4%) | 153 (18%) | 263 (11%) | 190 (10%) | 591 (13%) |
| Missing | 12 | 15 | 1,068 | 2 | 0 | 24 | 8 | 13 | 13 | 1 | 4 | 2 | 5 |
| tEPEC detected | 412 (27%) | 507 (25%) | 346 (24%) | 19 (18%) | 19 (13%) | 403 (18%) | 294 (26%) | 453 (21%) | 258 (24%) | 288 (34%) | 427 (18%) | 440 (24%) | 624 (14%) |
| Missing | 7 | 9 | 1,064 | 2 | 0 | 22 | 8 | 10 | 2 | 0 | 4 | 4 | 8 |
| *Campylobacter jejuni/coli* detected | 583 (38%) | 964 (47%) | 612 (42%) | 42 (40%) | 50 (34%) | 780 (34%) | 544 (48%) | 1,043 (49%) | 297 (28%) | 509 (60%) | 1,061 (45%) | 690 (37%) | 1,310 (29%) |
| Missing | 10 | 5 | 1,069 | 2 | 0 | 26 | 10 | 6 | 1 | 0 | 4 | 7 | 12 |
| Sapovirus detected | 214 (14%) | 237 (12%) | 162 (11%) | 24 (23%) | 30 (21%) | 392 (17%) | 145 (13%) | 284 (13%) | 127 (12%) | 198 (23%) | 237 (10%) | 228 (12%) | 414 (9.3%) |
| Missing | 12 | 18 | 1,069 | 2 | 0 | 20 | 7 | 13 | 11 | 1 | 3 | 22 | 74 |
| *Giardia* detected | 778 (52%) | 1,044 (51%) | 806 (56%) | 45 (42%) | 37 (25%) | 1,046 (46%) | 438 (53%) | 778 (47%) | 439 (59%) | 273 (32%) | 505 (21%) | 632 (34%) | 1,197 (27%) |
| Missing | 18 | 7 | 1,080 | 1 | 0 | 32 | 330 | 477 | 322 | 2 | 15 | 13 | 37 |
| *E. bieneusi* detected | 137 (9.0%) | 189 (9.2%) | 152 (10%) | 9 (8.5%) | 8 (5.5%) | 194 (8.5%) | 132 (12%) | 226 (11%) | 112 (10%) | 78 (9.2%) | 186 (7.9%) | 155 (8.3%) | 335 (7.4%) |
| Missing | 12 | 10 | 1,066 | 1 | 0 | 23 | 8 | 8 | 2 | 0 | 4 | 4 | 8 |
| EAEC detected | 962 (63%) | 1,097 (53%) | 901 (61%) | 55 (53%) | 81 (55%) | 1,290 (57%) | 752 (67%) | 1,219 (58%) | 675 (64%) | 629 (74%) | 1,472 (62%) | 1,189 (64%) | 2,363 (53%) |
| Missing | 9 | 11 | 1,063 | 3 | 0 | 24 | 34 | 37 | 17 | 0 | 4 | 25 | 41 |

[1]n (%); [2]Controls in MALED are periods without diarrhea among 2,220 children

**Figure S1.** Effects of *Shigella*-attributable diarrhea on height-for-age z-score in the following 60-90 days compared to diarrhea with no pathogen etiology identified in each study and in an individual patient data (IPD) meta-analysis. Estimates are of the controlled direct effects of *Shigella* when treated with guideline recommended antibiotics (blue), when treated with not recommended but possibly effective antibiotics (purple) and when treated with no or ineffective antibiotics (red).

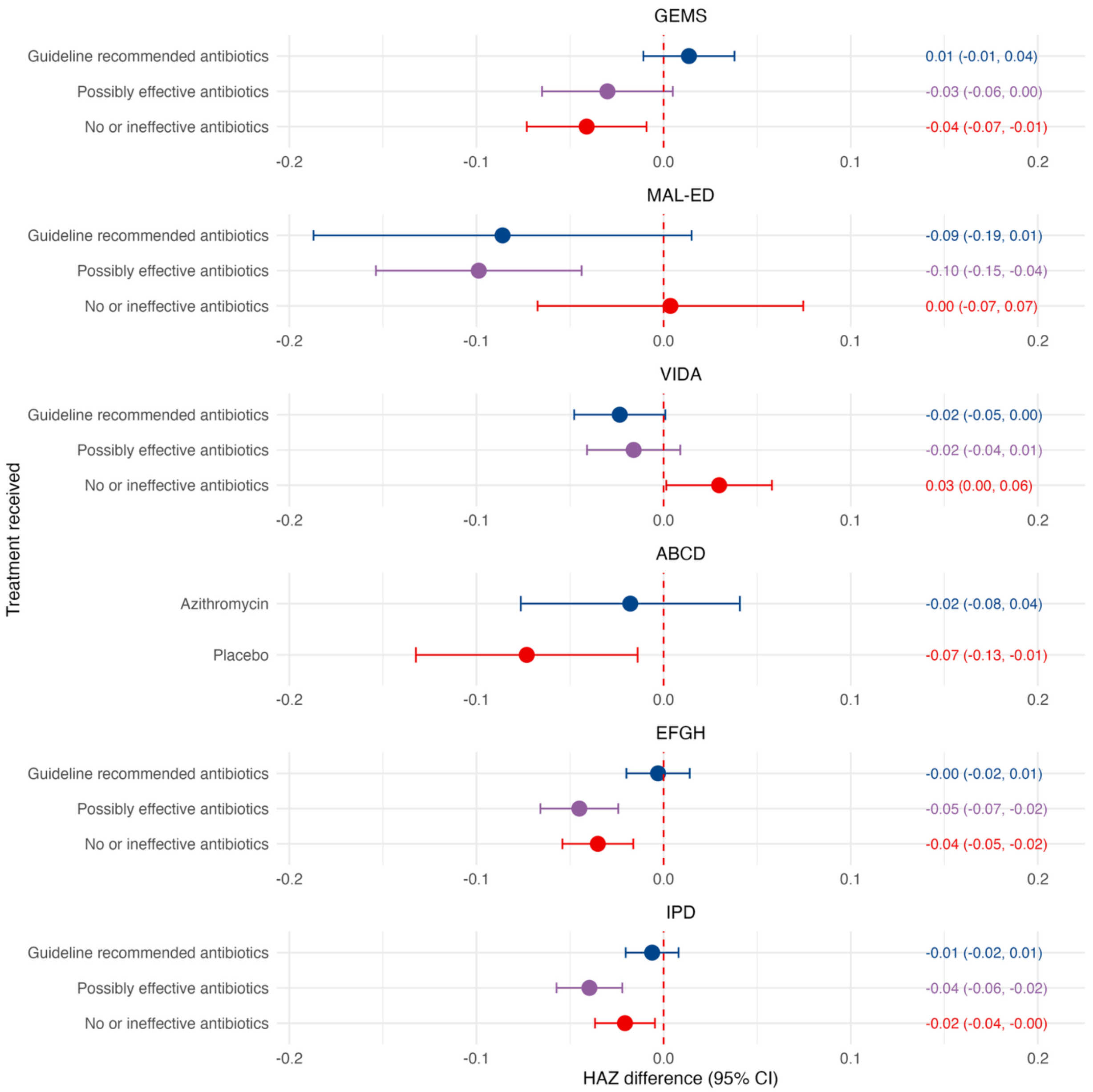

**Figure S2.** Study-specific heterogeneity by age in the effect of *Shigella*-attributable diarrhea on HAZ in the following 60-90 days compared to episodes with no pathogen etiology identified.

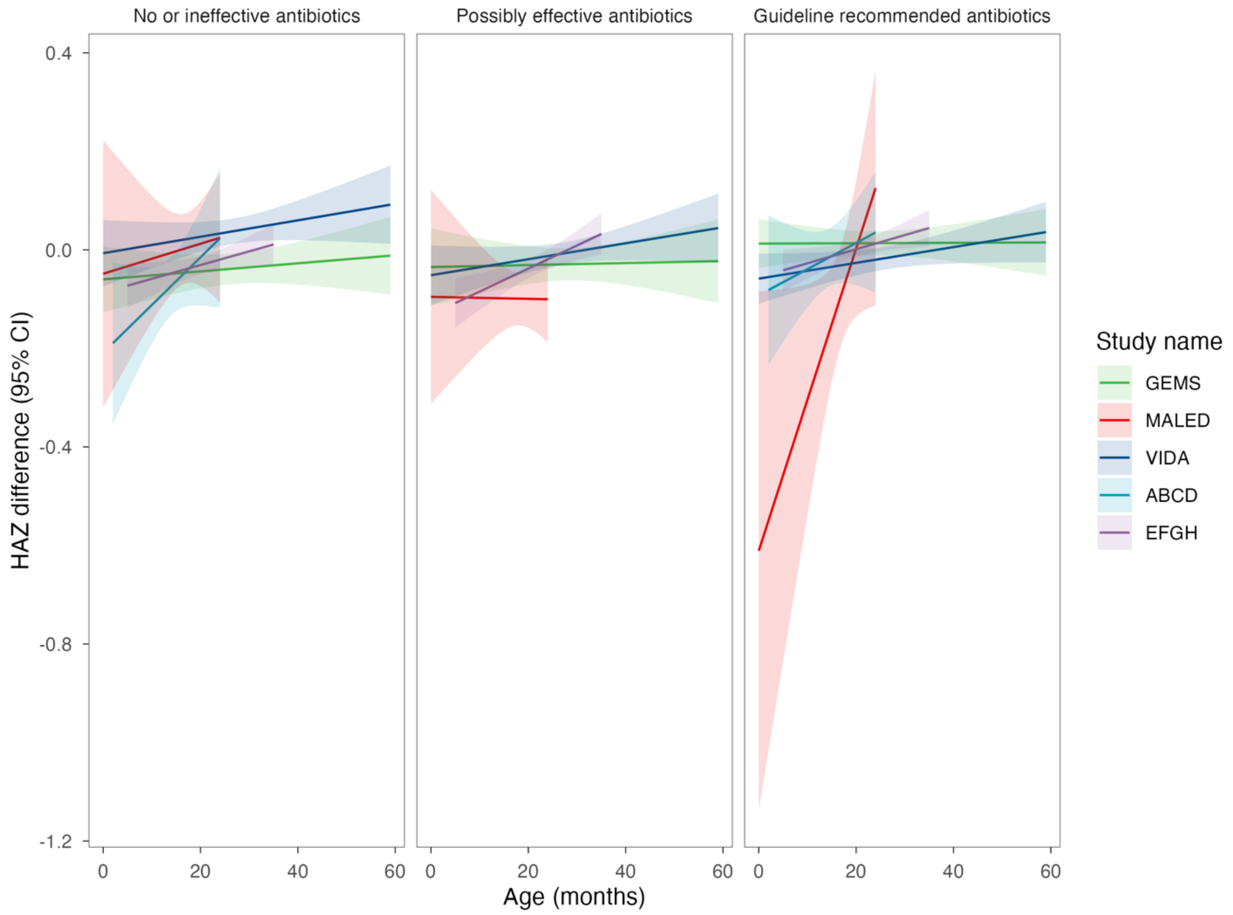

**Figure S3.** Effects of *Shigella*-attributable diarrhea on height-for-age z-score in the following 60-90 days with observed antibiotic use, compared to non-diarrheal controls in an individual patient data meta-analysis. Estimates are of the controlled direct effects of all *Shigella* episodes (purple), less severe diarrhea (blue), and moderate to severe diarrhea (red).

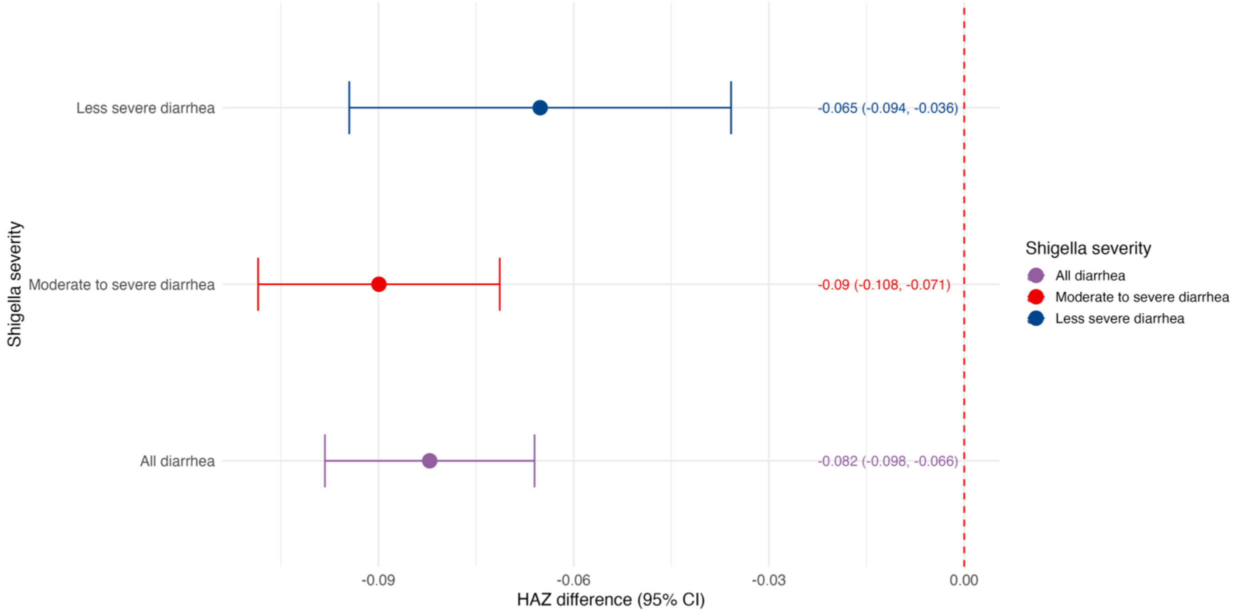

**Figure S4.** Effects of *Shigella*-attributable diarrhea on height-for-age z-score in the following 60-90 days compared to non-diarrheal controls in each study and in an individual patient data (IPD) meta-analysis. Estimates are of the controlled direct effects of *Shigella* when treated with guideline recommended antibiotics (blue), when treated with not recommended but possibly effective antibiotics (purple) and when treated with no or ineffective antibiotics (red).

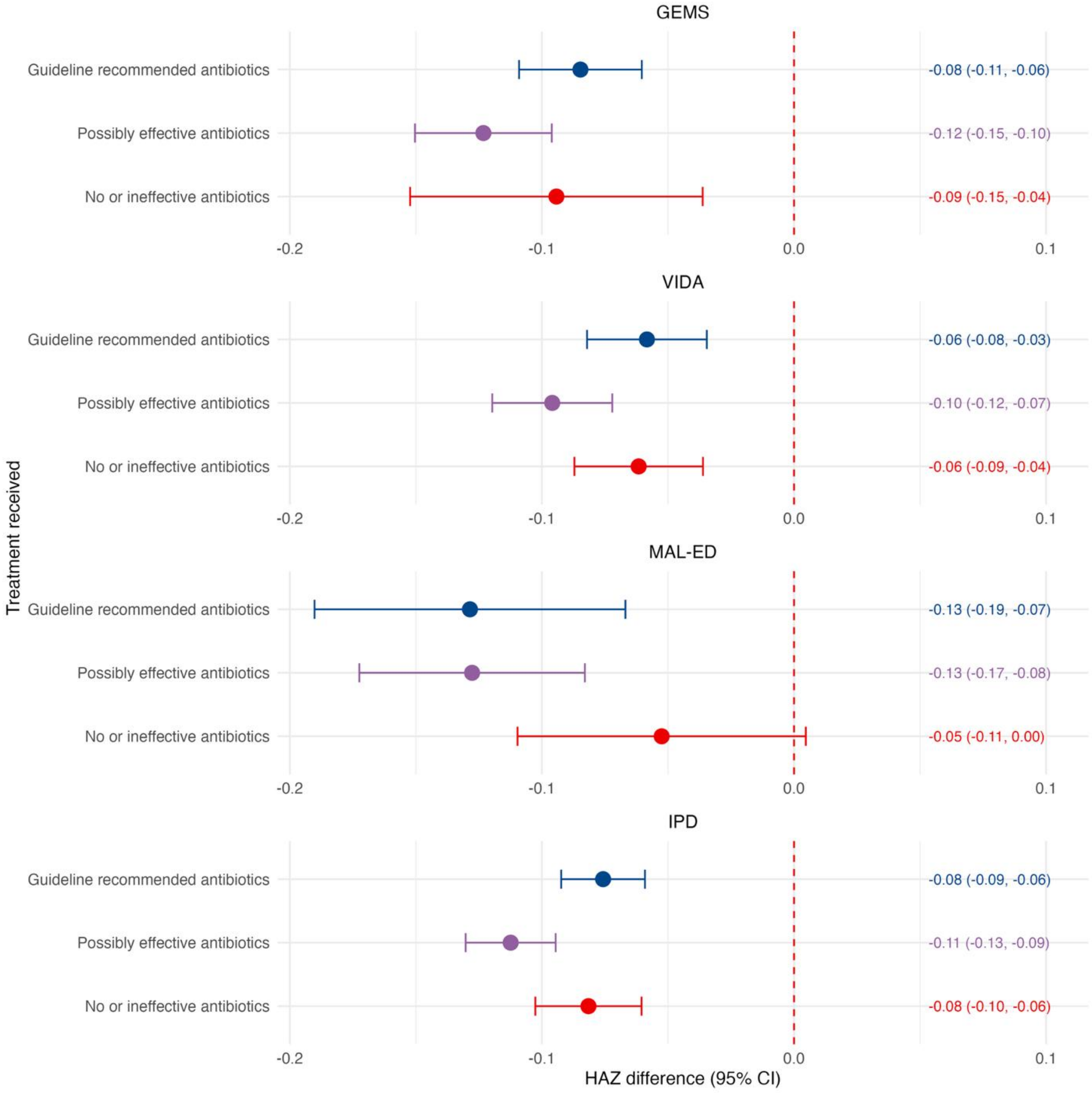

**Figure S5.** Effects of *Shigella*-attributable diarrhea on height-for-age z-score in the following 60-90 days with observed antibiotic use, compared to non-diarrheal controls in an individual patient data meta-analysis. Estimates are of the controlled direct effects of all *Shigella* episodes stratified by age 0-11 months (blue), age 12-23 months (purple), and age 24-59 months (red).

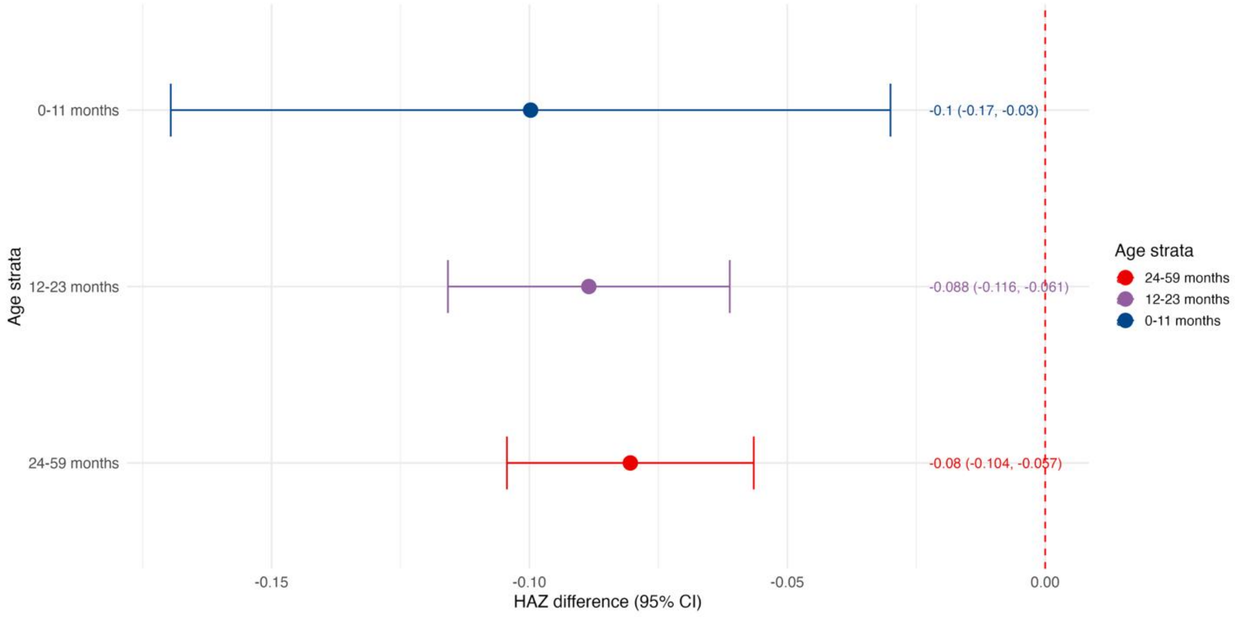

**Figure S6.** Study-specific heterogeneity by age in the effect of *Shigella*-attributable diarrhea on HAZ in the following 60-90 days compared to non-diarrheal controls.

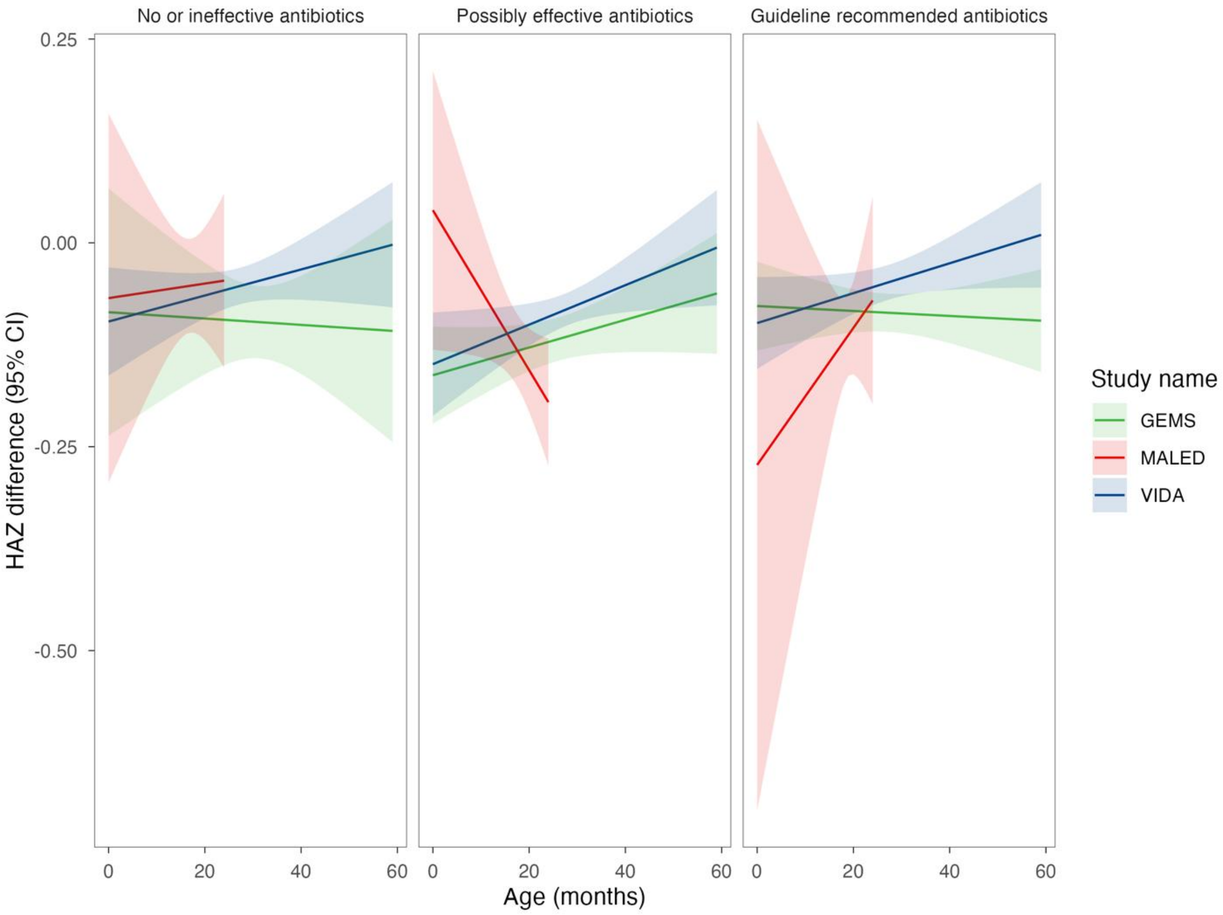

**Figure S7.** Effects of *Shigella*-attributable diarrhea, stratified by dysentery versus watery diarrhea (no dysentery), on height-for-age z-score in the following 60-90 days compared to diarrhea with no pathogen etiology in an individual patient data meta-analysis, stratified by age group. Estimates are of the controlled direct effects of *Shigella* when treated with guideline recommended antibiotics (blue), when treated with not recommended but possibly effective antibiotics (purple), and when treated with no antibiotics or ineffective antibiotics (red).

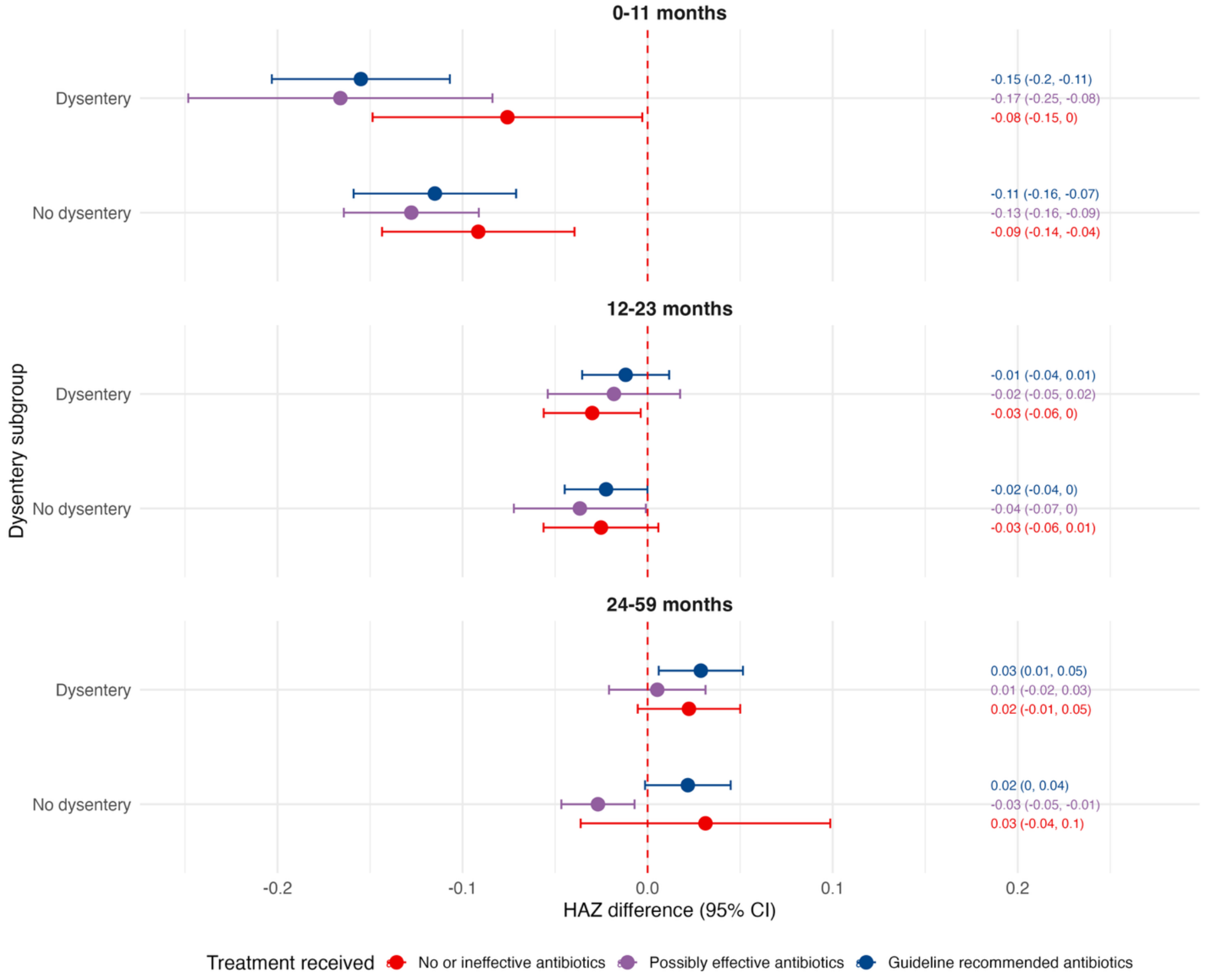

**Figure S8.** Effects of *Shigella*-attributable diarrhea, stratified by dysentery versus watery diarrhea (no dysentery), on height-for-age z-score in the following 60-90 days compared to non-diarrheal controls in an individual patient data meta-analysis, stratified by age group. Estimates are of the controlled direct effects of *Shigella* when treated with guideline recommended antibiotics (blue), when treated with not recommended but possibly effective antibiotics (purple), and when treated with no antibiotics or ineffective antibiotics (red).

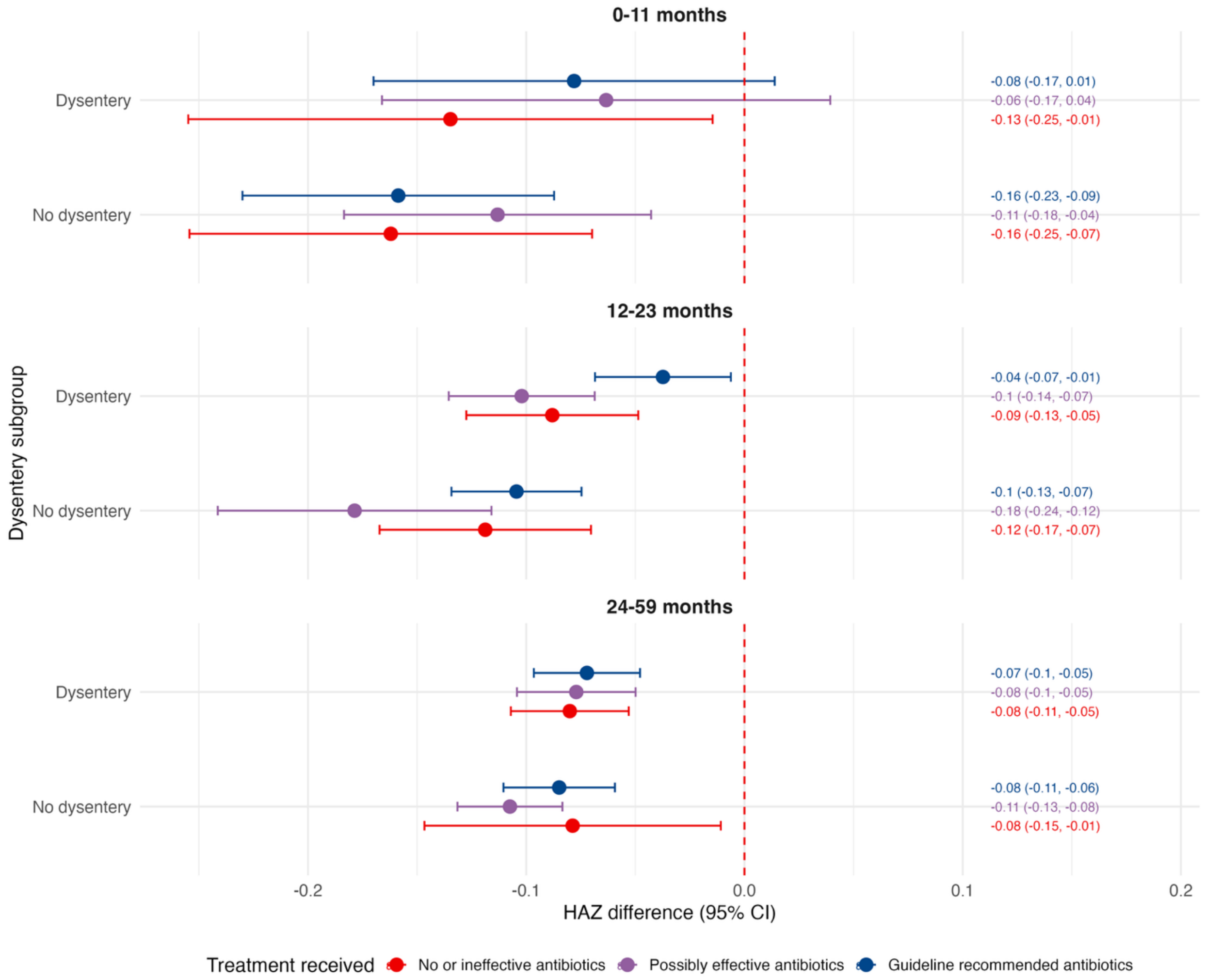

**Figure S9.** Effects of *Shigella*-attributable diarrhea, stratified by *Shigella* species, on height-for-age z-score in the following 60-90 days compared to diarrhea with no pathogen etiology in an individual patient data meta-analysis. Estimates are of the controlled direct effects of *Shigella* when treated with guideline recommended antibiotics (blue), when treated with not recommended but possibly effective antibiotics (purple), and when treated with no antibiotics or ineffective antibiotics (red).

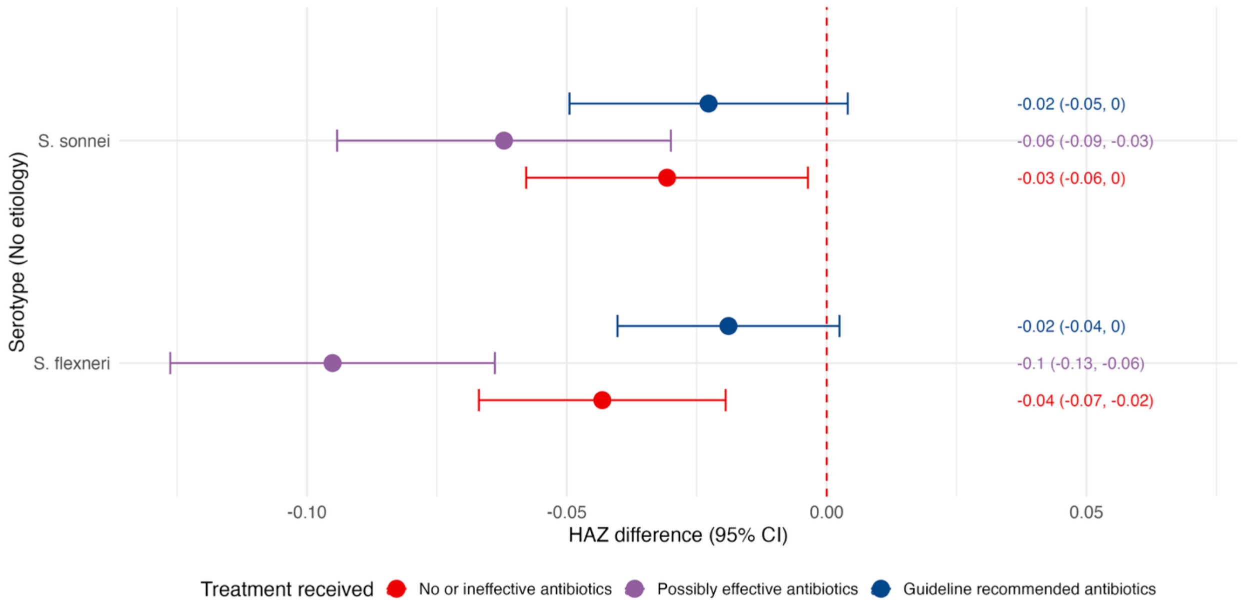

**Figure S10.** Effects of *Shigella*-attributable diarrhea, stratified by *Shigella* species, on height-for-age z-score in the following 60-90 days compared to non-diarrheal controls in an individual patient data meta-analysis. Estimates are of the controlled direct effects of *Shigella* when treated with guideline recommended antibiotics (blue), when treated with not recommended but possibly effective antibiotics (purple), and when treated with no antibiotics or ineffective antibiotics (red).

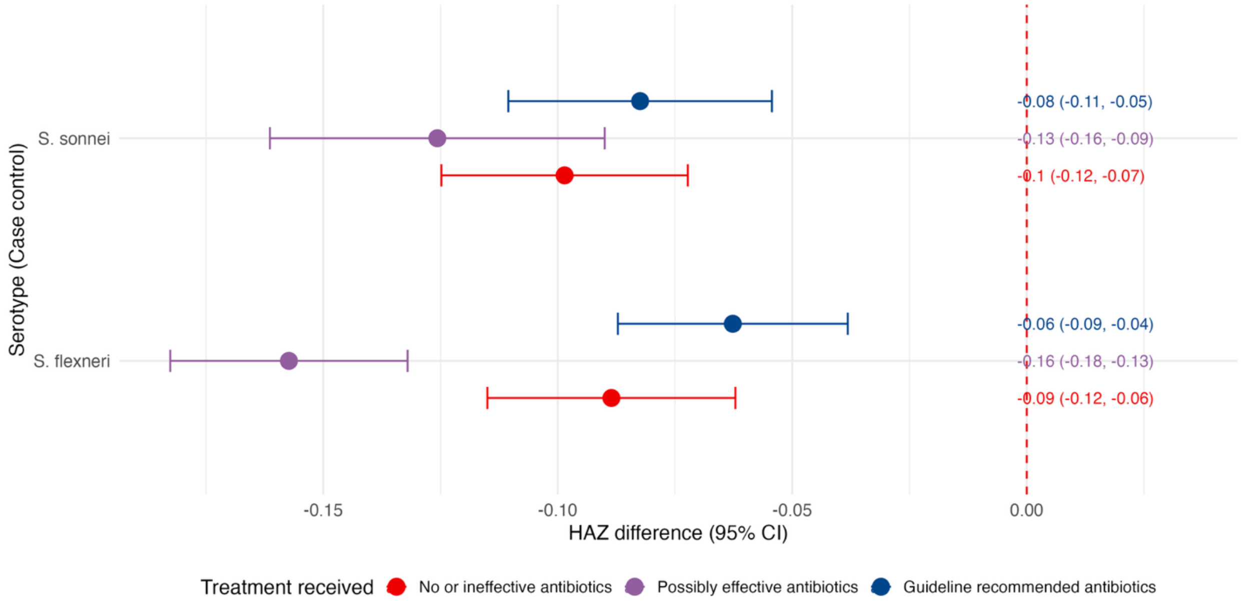

**Figure S11.** Effects of _Shigella_ culture-positive diarrhea on height-for-age z-score in the following 60-90 days compared to diarrhea with no pathogen etiology identified in each study and in an individual patient data meta-analysis. Estimates are of the controlled direct effects of *Shigella* when treated with guideline recommended antibiotics (blue), when treated with not recommended but possibly effective antibiotics (purple), and when treated with no antibiotics or ineffective antibiotics (red).

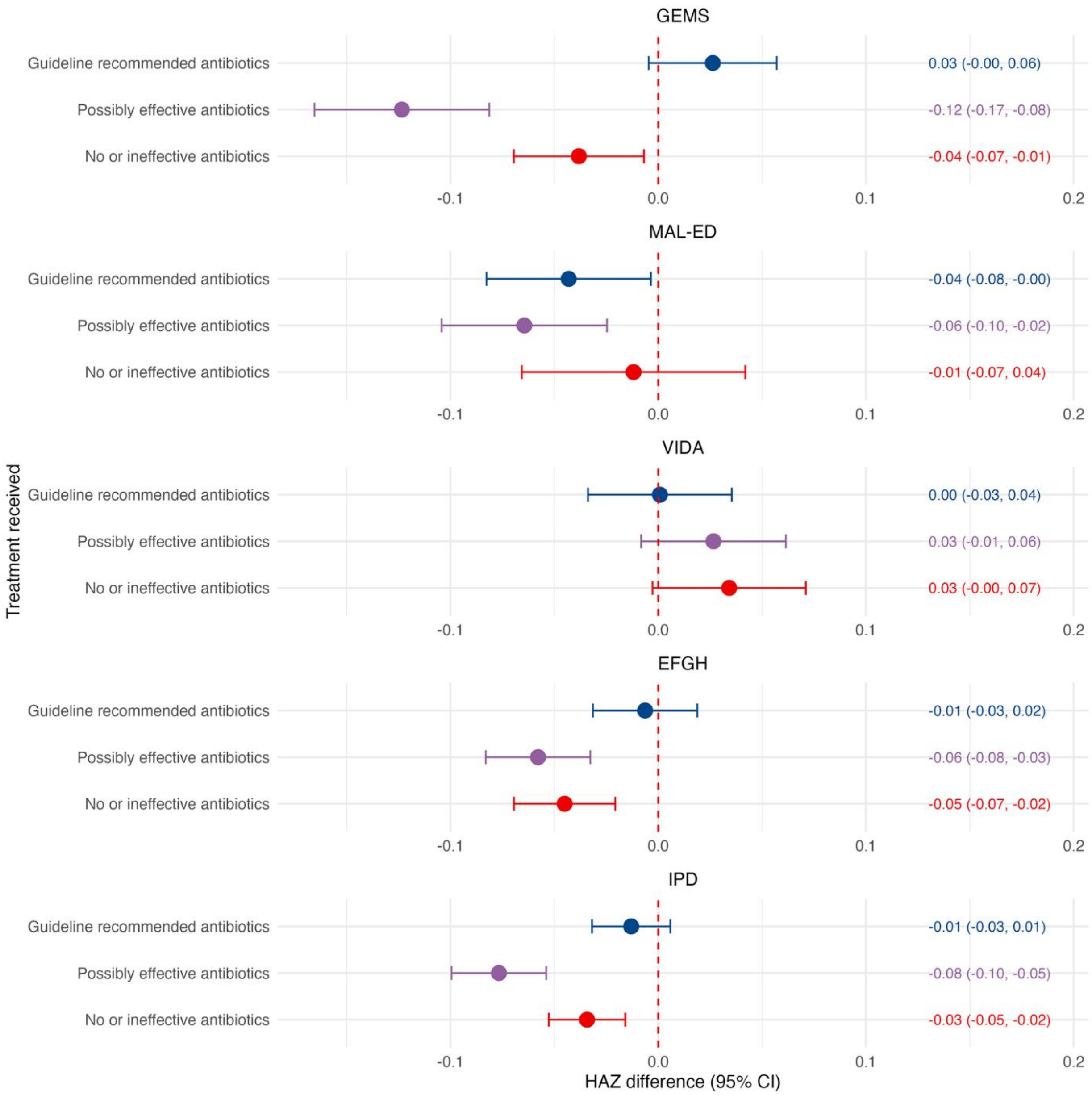

**Figure S12.** Effects of <u>*Shigella* culture-positive diarrhea</u> on height-for-age z-score in the following 60-90 days compared to non-diarrheal controls in each study and in an individual patient data meta-analysis. Estimates are of the controlled direct effects of *Shigella* when treated with guideline recommended antibiotics (blue), when treated with not recommended but possibly effective antibiotics (purple), and when treated with no antibiotics or ineffective antibiotics (red).

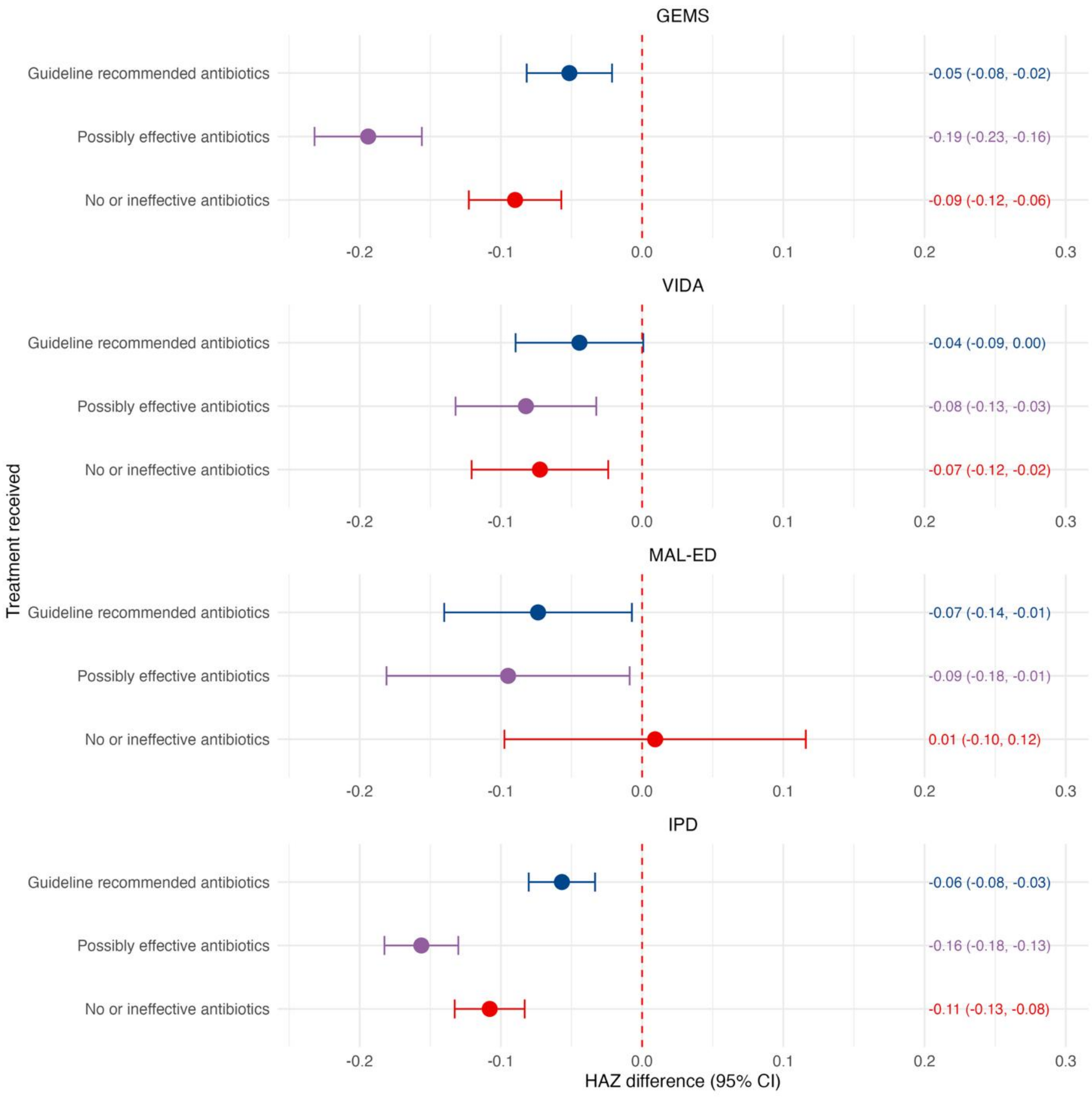

**Figure S13.** Effects of *Shigella*-attributable diarrhea on height-for-age z-score in the following 60-90 days <u>compared to all other diarrhea</u> identified in each study and in an individual patient data meta-analysis. Estimates are of the controlled direct effects of *Shigella* when treated with guideline recommended antibiotics (blue), when treated with not recommended but possibly effective antibiotics (purple), and when treated with no antibiotics or ineffective antibiotics (red).

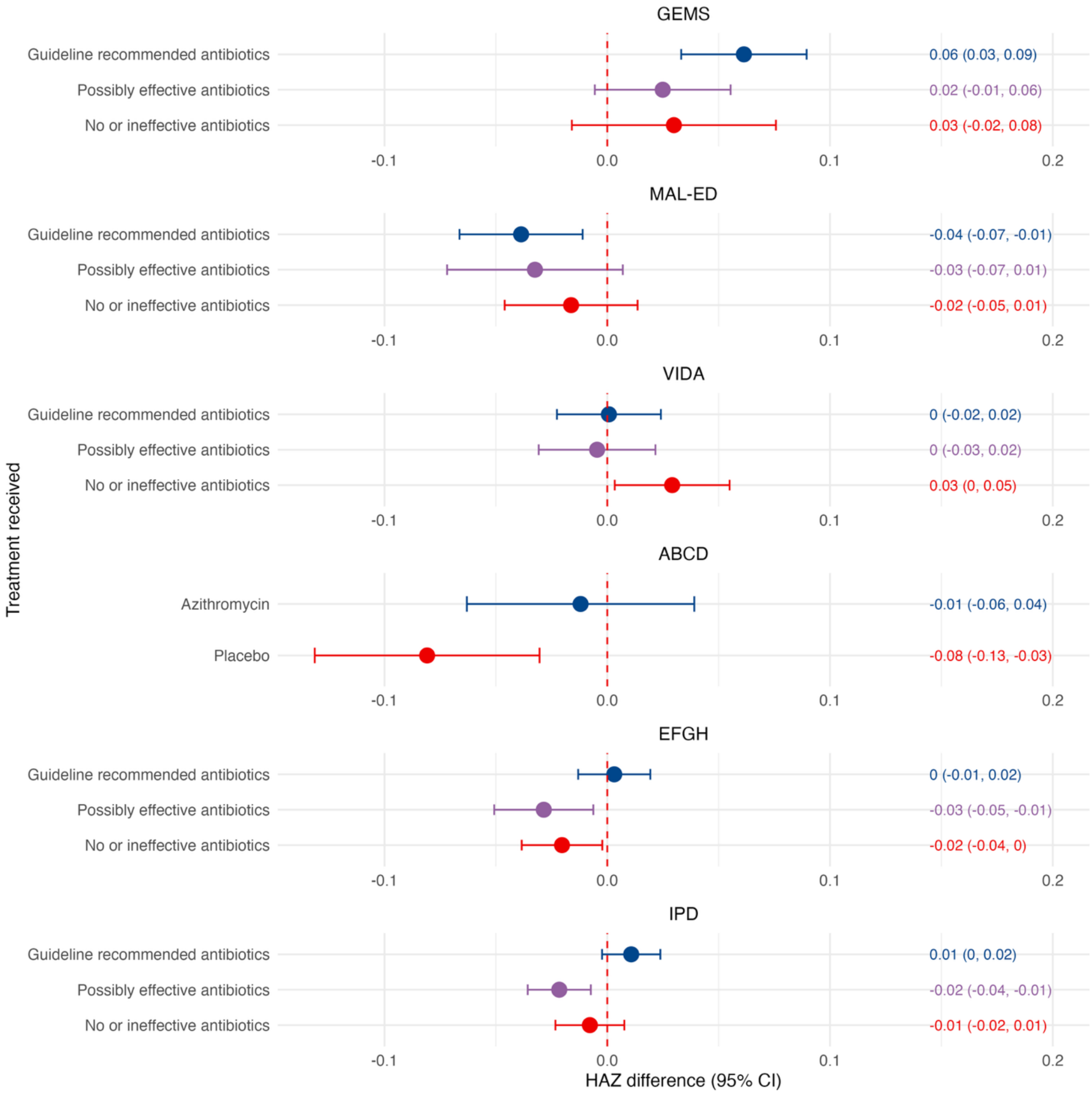

### References


1 Kotloff KL, Nataro JP, Blackwelder WC, *et al.* Burden and aetiology of diarrhoeal disease in infants and young children in developing countries (the Global Enteric Multicenter Study, GEMS): a prospective, case-control study. *The Lancet* May 20, 14. http://linkinghub.elsevier.com/retrieve/pii/S0140673613608442.

2 MAL-ED Network Investigators. The MAL-ED study: a multinational and multidisciplinary approach to understand the relationship between enteric pathogens, malnutrition, gut physiology, physical growth, cognitive development, and immune responses in infants and children up to 2 years of age in resource-poor environments. *Clin Infect Dis* 2014; **59 Suppl 4**: S193-206.

3 Richard SA, Barrett LJ, Guerrant RL, Checkley W, Miller MA, MAL-ED Network Investigators. Disease surveillance methods used in the 8-site MAL-ED cohort study. *Clin Infect Dis* 2014; **59 Suppl 4**: S220-224.

4 Platts-Mills JA, Babji S, Bodhidatta L, *et al.* Pathogen-specific burdens of community diarrhoea in developing countries: a multisite birth cohort study (MAL-ED). *Lancet Glob Health* 2015; **3**: e564-75.

5 Buchwald AG, Verani JR, Keita AM, *et al.* Etiology, Presentation, and Risk Factors for Diarrheal Syndromes in 3 Sub-Saharan African Countries After the Introduction of Rotavirus Vaccines From the Vaccine Impact on Diarrhea in Africa (VIDA) Study. *Clinical Infectious Diseases* 2023; **76**: S12–22.

6 ABCD study team. A double-blind placebo-controlled trial of azithromycin to reduce mortality and improve growth in high-risk young children with non-bloody diarrhoea in low resource settings: the Antibiotics for Children with Diarrhoea (ABCD) trial protocol. *Trials* 2020; **21**: 71.

7 Antibiotics for Children With Diarrhea (ABCD) Study Group, Ahmed T, Chisti MJ, *et al.* Effect of 3 Days of Oral Azithromycin on Young Children With Acute Diarrhea in Low-Resource Settings: A Randomized Clinical Trial. *JAMA Netw Open* 2021; **4**: e2136726.

8 Pavlinac PB, Platts-Mills JA, Liu J, *et al.* Azithromycin for Bacterial Watery Diarrhea: A Reanalysis of the AntiBiotics for Children With Severe Diarrhea (ABCD) Trial Incorporating Molecular Diagnostics. *J Infect Dis* 2024; **229**: 988–98.

9 van der Laan MJ, Polley EC, Hubbard AE. Super learner. *Stat Appl Genet Mol Biol* 2007; **6**: Article25.

10 Polley E, LeDell E, Kennedy C, Lendle S, Laan M van der. SuperLearner: Super Learner Prediction. 2025; published online Dec 21. https://cran.r-project.org/web/packages/SuperLearner/index.html (accessed June 23, 2026).

11 Gruber S, Phillips RV, Lee H, van der Laan MJ. Data-Adaptive Selection of the Propensity Score Truncation Level for Inverse-Probability-Weighted and Targeted Maximum Likelihood Estimators of Marginal Point Treatment Effects. *Am J Epidemiol* 2022; **191**: 1640–51.

12 Schnitzer ME, van der Laan MJ, Moodie EEM, Platt RW. EFFECT OF BREASTFEEDING ON GASTROINTESTINAL INFECTION IN INFANTS: A TARGETED MAXIMUM LIKELIHOOD APPROACH FOR CLUSTERED LONGITUDINAL DATA. *Ann Appl Stat* 2014; **8**: 703–25.

13 Rosenblum M, Laan MJ van der. Targeted Maximum Likelihood Estimation of the Parameter of a Marginal Structural Model. *The International Journal of Biostatistics* 2010; **6**. DOI:10.2202/1557-4679.1238.

14 Neugebauer R, van der Laan M. Nonparametric causal effects based on marginal structural models. *Journal of Statistical Planning and Inference* 2007; **137**: 419–34.

15 Horne B, Badji H, Bhuiyan MTR, *et al.* Microbiological Methods Used in the Enterics for Global Health Shigella Surveillance Study. *Open Forum Infectious Diseases* 2024; **11**: S25–33.
